# Severe obesity may be an oligogenic condition: evidence from 1,714 adults seeking treatment in the UK National Health Service

**DOI:** 10.1101/2023.08.04.23293229

**Authors:** Almansoori Sumaya, Hasnat A Amin, Suzanne I. Alsters, Dale Handley, Andrianos M Yiorkas, Nikman Adli Nor Hashim, Nurul Hanis Ramzi, Sanjay Agrawal, Gianluca Bonaomi, Javed Ahmed, Peter Small, Sanjay Purkayastha, Mieke van Haelst, Robin G. Walters, Carel W le Roux, Harvinder S. Chahal, Fotios Drenos, Alexandra I Blakemore

## Abstract

Severe (class III) obesity is a chronic, relapsing condition, with a high burden of co-morbidity and mortality. Previous evidence has established genetics as an important contributing factor. We therefore designed a custom genotyping array to screen a cohort of UK patients seeking treatment for severe obesity. 1,714 participants were genotyped using a custom AXIOM array, focusing on rare (minor allele frequency <0.01) variants, with CADD-PHRED ≥15 in 78 genes known/suspected to cause Mendelian forms of obesity. Concordance analyses of 22 duplicate samples and 66 samples with whole exome sequence data revealed good genotyping reliability. We identified the proportion of study participants who carried, or were homozygous for, rare, predicted-deleterious variants in genes with dominant and recessive modes of inheritance (MOI), respectively. 27% of patients carried relevant mutations consistent with the expected MOI, which was very similar to the rate observed in the UKB 50K whole exome sequencing dataset. However, the clinical obesity cohort were more likely to carry two or more such variants, in separate genes, than UK Biobank participants (p = 0.018). In conclusion, our results provide evidence: that (i) custom genotyping arrays, used with improved algorithms can allow reliable, cost-effective screening for rare genetic variants; (ii) rare mutations in “obesity genes” may be at high prevalence among bariatric patients, as well as in the general population; and (iii) that severe obesity may have an oligogenic pattern of inheritance in some cases.

## INTRODUCTION

Severe (class III) obesity is a chronic, relapsing condition, with a high burden of co-morbidity and mortality [1]. It has been estimated that the healthcare costs for an individual with class III obesity are about 40% higher than for an individual who is of normal weight [2]. It is, therefore, essential to better understand aetiology of this disease.

Obesity is a complex heterogeneous disorder, with interacting environmental and genetic components. Genetic research has revealed the influence of common genetic variants with subtle effects on body mass index (BMI) or risk of obesity [3], but also the existence of monogenic and syndromic forms of obesity which are rare in general populations, but may be much more frequent in clinical obesity cohorts [4, 5]. Although there have been long-standing efforts directed at understanding the genetic underpinning of complex and severe forms of childhood obesity [6], most adults – even when very severely affected – remain un-investigated so that opportunities for precision medicine are limited. Additionally, despite mounting appreciation of the importance of oligogenic inheritance in other diseases [7-9], this form of inheritance has not been systematically explored in severe obesity.

Here we present an investigation aimed at exploring the prevalence of rare genetic forms of obesity in an UK-based clinical obesity cohort. For this work, we designed a custom genotyping array, including rare predicted-deleterious variants in relevant genes, and applied this to screening 1,714 unrelated adults seeking NHS treatment for severe obesity. We compared the results from this clinical cohort to the 50K exome sequencing dataset from the UK Biobank. Based on our previous findings [10], we also explored these datasets for evidence of oligogenic inheritance in severe obesity.

## METHODS

### Cohort descriptions

### Personalised Medicine for Morbid Obesity

A total of 2,556 unrelated adults were recruited between 2011 and 2021 for the Personalised Medicine for Morbid Obesity (PMMO) NHS-registered portfolio trial (NCT01365416;[11]). Participants had a BMI ≥40 or a BMI ≥35 with significant co-morbidities. Of the 2,556 participants, 1,469 were recruited prospectively across different hospital sites (The Imperial College NHS Weight Centre, London, United Kingdom; Chelsea and Westminster Hospital, London, United Kingdom; and Royal Derbyshire Hospital, Derby, United Kingdom, City Hospitals Sunderland NHS Foundation Trust, United Kingdom, Charing Cross Hospital, London, United Kingdom) and 1,087 were recruited retrospectively using NHS electronic GP records.

Ethical approval for the collection and use of data for the PMMO study was given by the NRES Committee London Riverside (REC reference 11\LO\0935) and the research was performed in accordance with the principles of the Declaration of Helsinki. All participants gave informed consent.

### UK Biobank

The UK Biobank (UKB) is a large prospective cohort study of approximately 500,000 participants, recruited between 2006 and 2011. The 22 UKB assessment centres, located throughout England, Wales and Scotland, collected baseline data from the participants in the form of questionnaires, physical and cognitive tests and blood and urine samples [12]. The age range of the participants at the time of enrolment in the study was between 40 and 69 years of age. Men represent just under 46% of the sample. Permission to use UKB data for the investigation of obesity-related traits is provided under UKB application number 62265.

### Design of a custom genotyping array

A customised genotyping array was created which examines genes related to Mendelian forms of obesity and diabetes (as well as other conditions beyond the scope of this particular study). For the purpose of this study, 27 genes from an NHS panel typically used for clinical indication of severe early-onset obesity [13], as well as 51 genes related to syndromic and non-syndromic obesity, were utilised (Supplementary Table 1).

Selection of rare variants: all variants in HGMD (Human Gene Mutation Database) at each of the selected genes were selected irrespective of their phenotype or degree of pathogenicity. Additionally, gnomAD v2.1.1 was used to select variants that were located at the selected genes and had a MAF of <0.01. Due to the space limitations of the array, it was necessary to further prioritise variants that were identified from gnomAD: only missense, frameshift and stop-gain variants were included; and missense variants were filtered further using SIFT (damaging), PolyPhen (probably damaging) and CADD (CADD-PHRED >15) [14, 15].

There are several known gain-of-function variants in the *MC4R* gene [16] that are associated with leanness. These were excluded from the analyses, except where stated otherwise.

### Genotyping and quality control

### PMMO

Genomic DNA was prepared from either blood or saliva samples using standard methods. In total, 1,766 samples from the PMMO cohort underwent genotyping. Genotyping was performed using a custom Axiom genotyping array (see previous section) prepared by ThermoFisher and was performed according to ThermoFisher guidelines [17] by Oxford Genomics, and processed by the GeneTitan Multi-channel instrument. Probe clustering was performed using the AxiomGT1 algorithm in the Axiom analysis suite V5.1.1 and variants were classified into three categories based on their clustering pattern: MonoHighResolution means that the variant is monomorphic; NoMinorHom means that no participants were homozygous for the alternative allele; and PolyHighResolution indicates the presence of both heterozygotes and homozygotes for the alternative allele. Rare heterozygosity adjustment was used to adjust for multi-probe mismatches, which substantially improves correct rare variant calls for very rare variants [18]. Initial genotyping filtering was conducted according to the Axiom array guidelines, with the DQC threshold set to 82%.

Samples were removed from further analyses if: they did not pass the QC metrics provided in the previous paragraph (n = 25); and/or they were duplicates, included for QC (n = 22); and/or they showed a high degree of heterozygosity or relatedness (n = 0); and/or they displayed discordant genetic and self-reported sex (n = 5). 1,714 samples remained after QC.

Whole exome sequencing (WES) data was available for 66 of these participants. Further details of how this was performed are available elsewhere [19].

### UK Biobank

For the 50k whole exome sequencing (WES) release from the UK Biobank (UKB), 49,960 samples had undergone exome capture using the IDT xGen Exome Research Panel v1.0 and sequenced using the Illumina Novaseq 6000 platform. Variant and sample QC was pre-performed by UKB and is described elsewhere [20]. Additionally, only variants that were both included on our custom array and located within the regions targeted by the IDT xGen Exome Research Panel v1.0 [21] were used for any analyses that used UKB data. Lastly, for all related pairs in the WES dataset, one was removed at random.

### Annotation

Variants were annotated using Variant Effect Predictor [22], with the annotations associated with the canonical transcript being selected if a variant was associated with more than one transcript. Variants were also annotated with CADD [23] and LOFTEE [24]. Variants were not considered for further analyses if they were exclusively annotated as being intronic variants and/or if they had a CADD-PHRED score of <15 and/or if they were low confidence variants according to LOFTEE.

### Inspection of cluster plots

We also manually inspected the cluster plots for variants that had at least one carrier. We found several variants that had an unexpectedly high MAF in our cohort and chose to exclude some of these variants due to the cluster plots for different probes targeting the same variant being inconsistent, or due to the presence of a common variant nearby which we believed may have interfered with the assay. Please see Appendix 1 for details of which variants were excluded and why.

### Statistical Analyses

All statistical analyses were carried out using R 4.2.1 [25], unless otherwise stated. PLINK 2.0 [26] was used to calculate per-sample concordance. Fisher’s exact test was used to compare proportions. The 95% confidence intervals used to plot error bars were calculated using 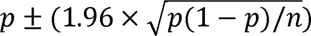, where *p* is the proportion and *n* is the sample size. The probability of an individual carrying a variant in a pair of genes was calculated by multiplying the proportion of individuals carrying at least one variant in one gene in that pair by the proportion of individuals carrying at least one variant in the other gene in that pair (individuals carrying more than one variant in the same gene were considered as having a genetic load of 1 – this conservative approach was adopted because we do not have phasing data that would allow us to determine cases of compound heterozygosity). We used a binomial exact test to test whether there is a statistically significant difference between the calculated probability and the actual probability.

## RESULTS

### Reproducibility of rare variant genotyping

We checked the concordance of genotypes between the 22 duplicate samples: the samples had an average concordance of 99.7% across all variants that passed QC. We next checked the concordance for PolyHighResolution (heterozygotes and homozygotes present for the alternative allele), NoMinorHom (no homozygotes for the alternative allele) and MonoHighResolution (monomorphic) variants separately: samples had an average concordance rate of 99.1%, 99.5% and 99.7%, respectively.

We next compared the genotypes for 66 samples for which we had whole exome sequence (WES) data. We found that the average concordance rate between genotyped samples and the WES samples across 1131 variants for which had WES data was 96.1%. The concordance for PolyHighResolution, NoMinorHom and MonoHighResolution variants was 94.0%, 98.8% and 95.2%, respectively.

### Summary of variants identified

Out of the 14887 variants that remained after filtering, 593 were carried by one or more PMMO participants, of which 359 were carried by a single participant. Of the 593 variants, 519 were missense variants, 29 were frameshift variants and 16 were stop-gain variants (Supplementary Table 2). 13784 out of 14887 variants had a CADD-PHRED score of 20 or higher (i.e. predicted to be in top 1% of all variants for deleteriousness): 530 of these were carried by one or more PMMO participants and 222 of these were carried by PMMO participants in accordance with their expected mode of inheritance (MOI), i.e. one copy was present for genes with dominant expected MOI, and they were in homozygous condition where the expected MOI was recessive (Supplementary Table 3).

A total of 6618 of 14887 variants were assigned to genes that are on an NHS panel used for clinical indication of severe early-onset obesity [13], of which 263 were carried by one or more PMMO participants. The remaining variants were from our extended obesity gene list, of which 330 were carried by one or more PMMO participants.

*MC4R* is possibly the best-investigated obesity gene; 15 PMMO participants (0.88%) carried variants in the *MC4R* gene, all in heterozygous (Supplementary Table 4). We note separately that 45 PMMO participants (2.6%) carried the *MC4R* rs2229616 gain-of-function variant (in contrast to 4% of UKB participants, p = 0.0026), but these are not included in our analyses.

### Prevalence of putatively Mendelian forms of obesity in the PMMO

We categorised our sample into three groups based on carrier status. Group 1 are individuals who do not carry any rare variants in obesity-implicated genes present on our array, group 2 are individuals who are heterozygous for rare variants in genes with a likely autosomal recessive mode of inheritance and the remaining individuals are in group 3 (i.e. individuals who are either homozygous for one or more rare variants with a recessive MOI, and/or heterozygous for rare variants in genes with a likely autosomal dominant MOI). Using a CADD-PHRED cut-off of 15 and the expanded gene list, 27.0% of the sample were categorised as group 3, indicating that they should be clinically evaluated for a possible Mendelian form of obesity. When considering only rare variants linked to genes that are on the NHS panel used for clinical indication of severe early-onset obesity [13], 8.6% of the samples are in group 3 (Figure 1).

**Figure 1.**
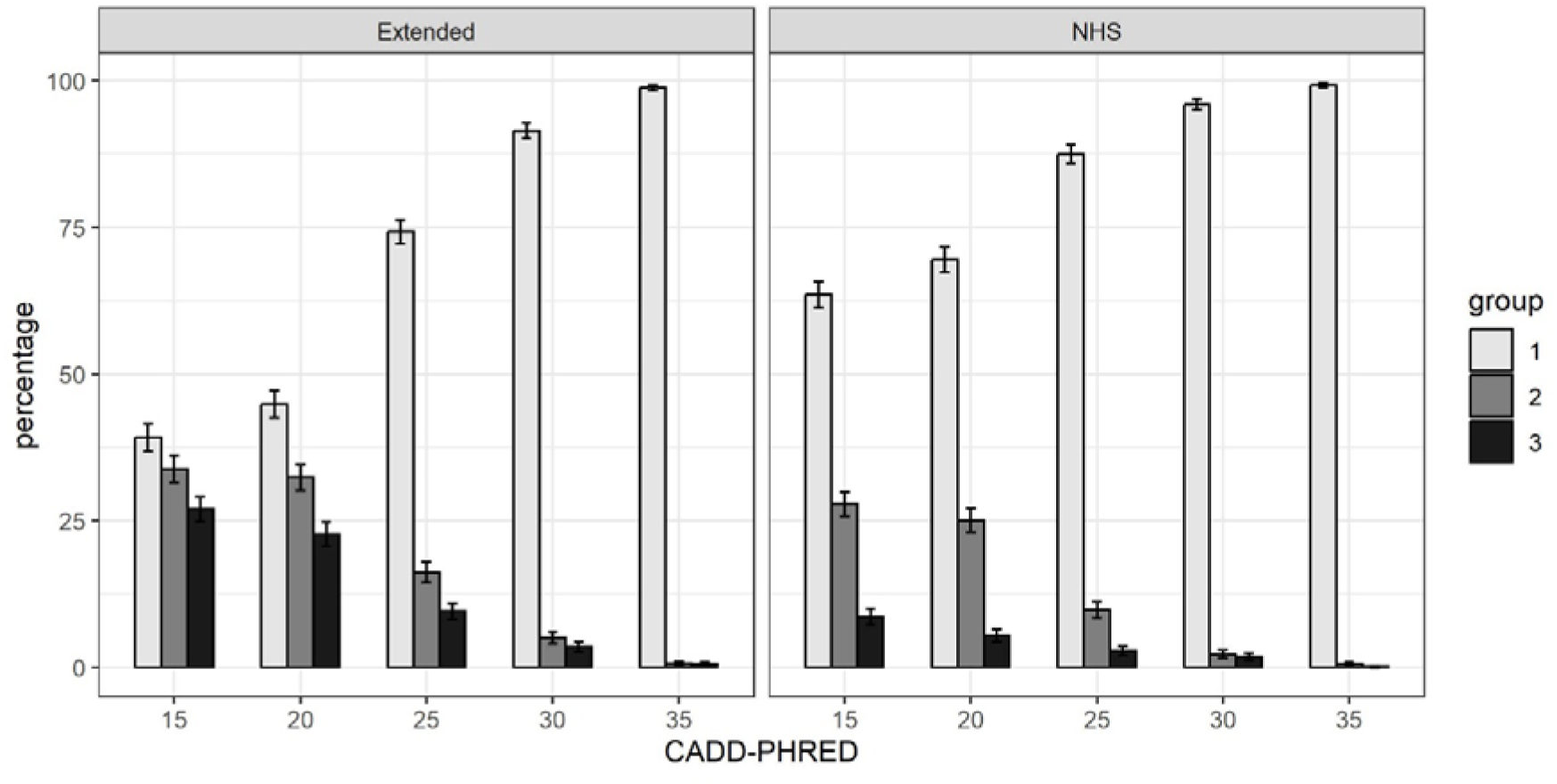
Percentage of PMMO participants that: (group 1) carry no relevant variants; (group 2) are heterozygous for variants in genes with likely recessive mode of inheritance; or (group 3) carry variants in heterozygous/homozygous state consistent with the expected mode of inheritance for that gene. The left panel shows the results for the extended gene list and the right panel shows results only for genes on the NHS list used for a clinical indication of severe early-onset obesity. The x-axis shows the CADD-PHRED score, which is an indication of likely deleteriousness, and variants with a CADD-PHRED ≥20 are among the top 1% most deleterious of mutations. Error bars represent 95% confidence intervals.

As expected, increasing the CADD-PHRED score threshold reduced the percentage of individuals who are in group 3: at a CADD-PHRED score of 25, 9.6% of participants had a suspected Mendelian form of obesity using the extended gene list (2.8% if considering only variants in genes on the NHS list for severe early-onset obesity).

### Comparison to carriage rates in a general population sample (UK Biobank)

We next compared the prevalence of putatively Mendelian forms of obesity in the PMMO to a general population sample – namely the UK Biobank (UKB). 24.5% of UKB participants are in group 3 (Figure 2), in comparison to 25.5% of PMMO participants (p = 0.33). Using only variants linked to genes on the NHS panel, there is no evidence that the proportion of group 3 participants differs between the PMMO and the UKB (8.3% vs. 7.9%, p = 0.55).

**Figure 2.**
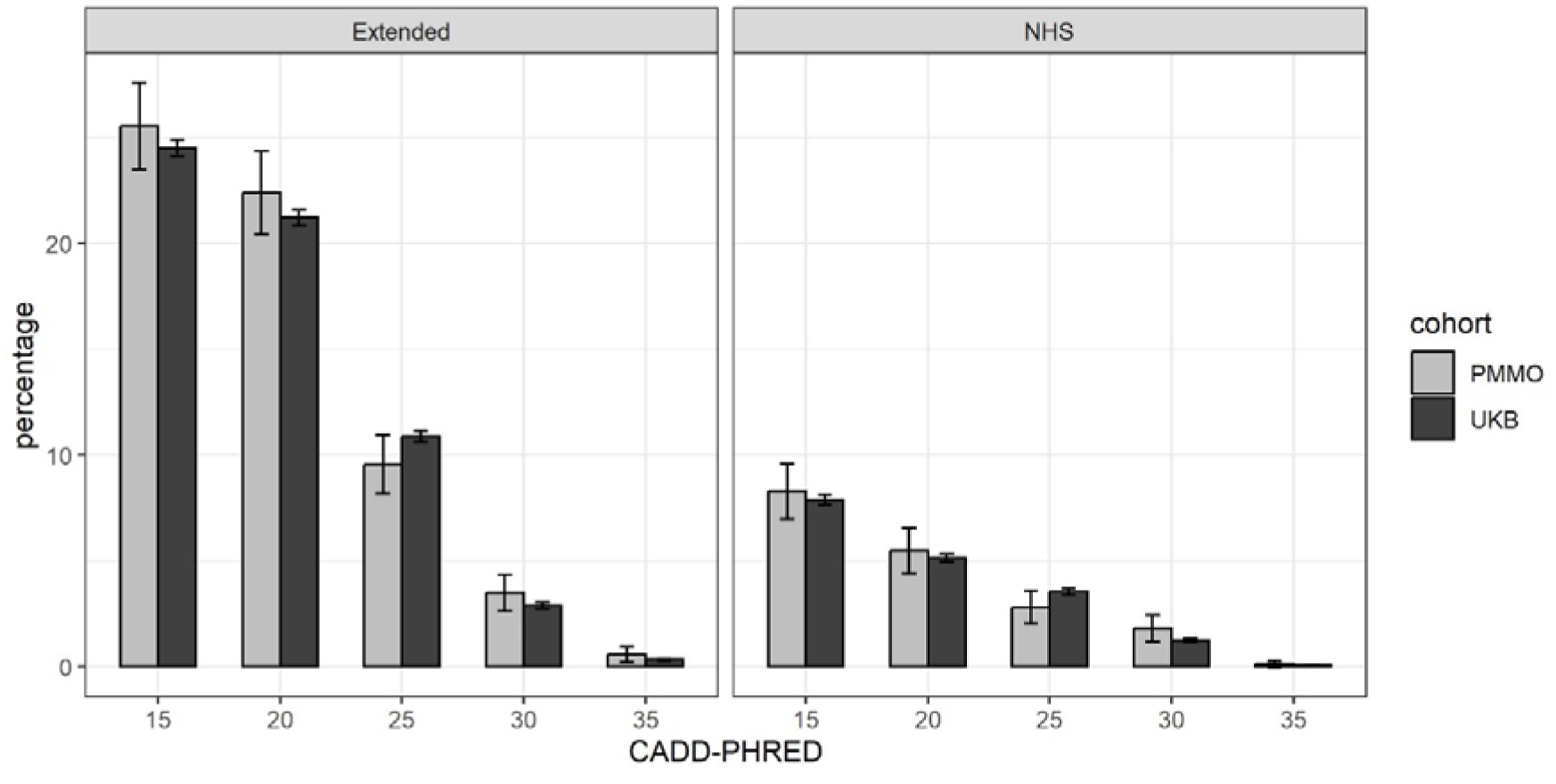
Percentage of PMMO and UK Biobank (UKB) participants that carry variants in heterozygous/homozygous state consistent with the expected mode of inheritance for that gene. The left panel shows the results for the extended gene list and the right panel shows results only for genes on the NHS list used for a clinical indication of severe early-onset obesity. The x-axis shows the CADD-PHRED score, which is an indication of likely deleteriousness, and variants with a CADD-PHRED ≥20 are among the top 1% most deleterious of mutations. Error bars represent 95% confidence intervals.

### Evidence for potentially oligogenic forms of obesity

A number of PMMO participants had variants (consistent with MOI) in multiple genes. We explored the possibility of oligogenic inheritance by comparing the proportion of group 3 PMMO participants who carried mutations in more than one gene (at CADD-PHRED score ≥15) with the proportion of group 3 UKB participants who carried mutations in more than one of those genes. 17.1% (N=75) of group 3 PMMO participants carry variants in two or more obesity-implicated genes, whereas this is the case for only 13.1% of group 3 UKB participants (p = 0.018). Please see Figure 3 for a visual representation of these results.

**Figure 3.**
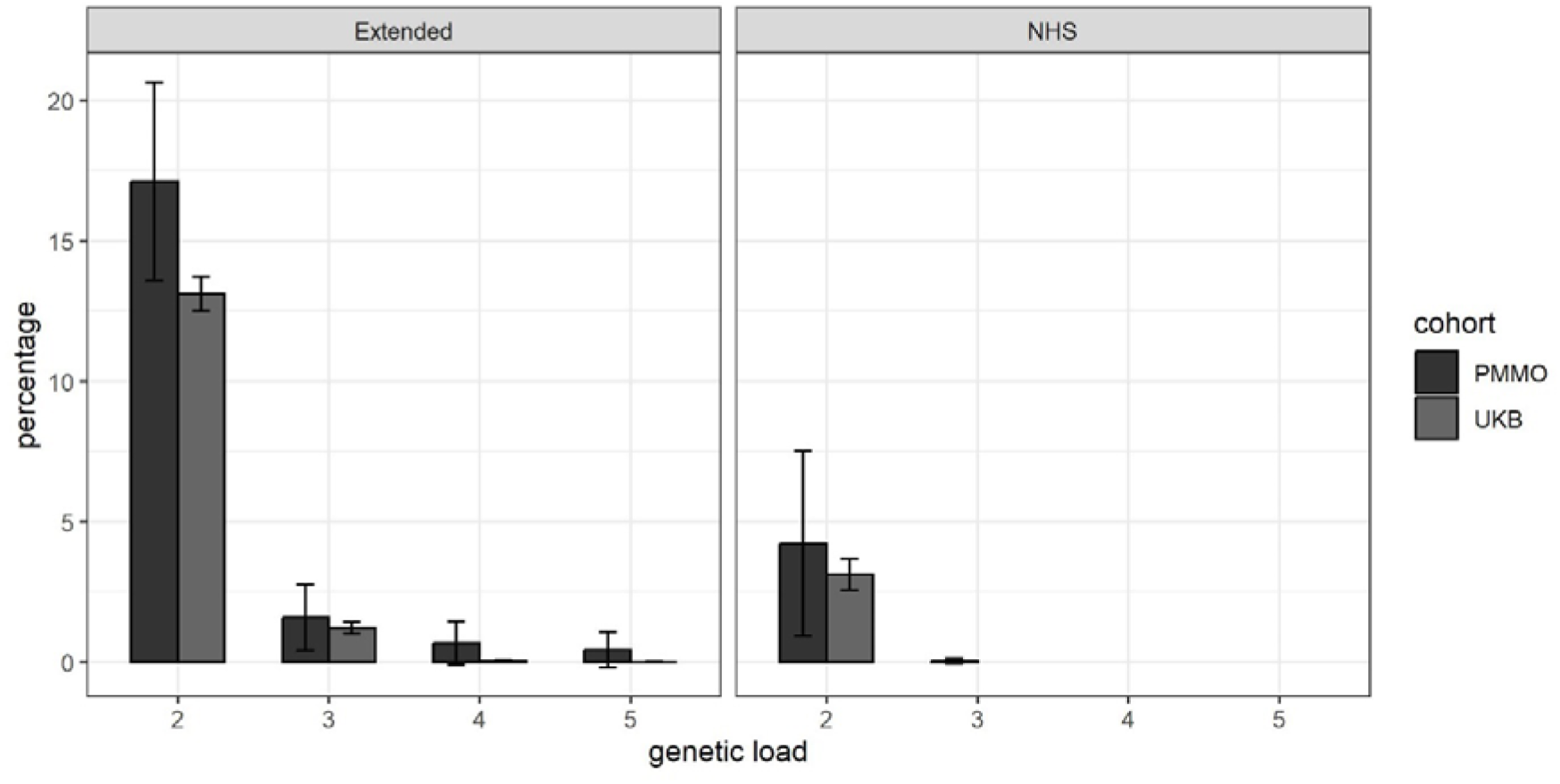
Percentage of PMMO and UK Biobank (UKB) participants that carry variants (in heterozygous/homozygous stated consistent with the expected mode of inheritance for that gene) in two or more genes. The left panel shows the results for the extended gene list and the right panel shows results only for genes on the NHS list used for a clinical indication of severe early-onset obesity. Error bars represent 95% confidence intervals.

We then explored whether PMMO participants in group 3 tended to carry variants in particular combinations of genes – displayed as a network plot in Figure 4. Since it is possible for individuals to carry variants in particular combinations of genes by chance, we calculated the probability for variants co-occurring in all combinations of genes shown in Figure 4 (see Supplementary Table 5) and found that the percentage of PMMO participants that carry one or more variants in both the *GSHR* and *MYT1L* genes is higher than would be expected by chance (0.46% vs. 0.013%, p = 0.0014). Please note that only PMMO participants who carry one or more variants according to the mode of inheritance (i.e. group 3) are considered for this analysis.

**Figure 4.**
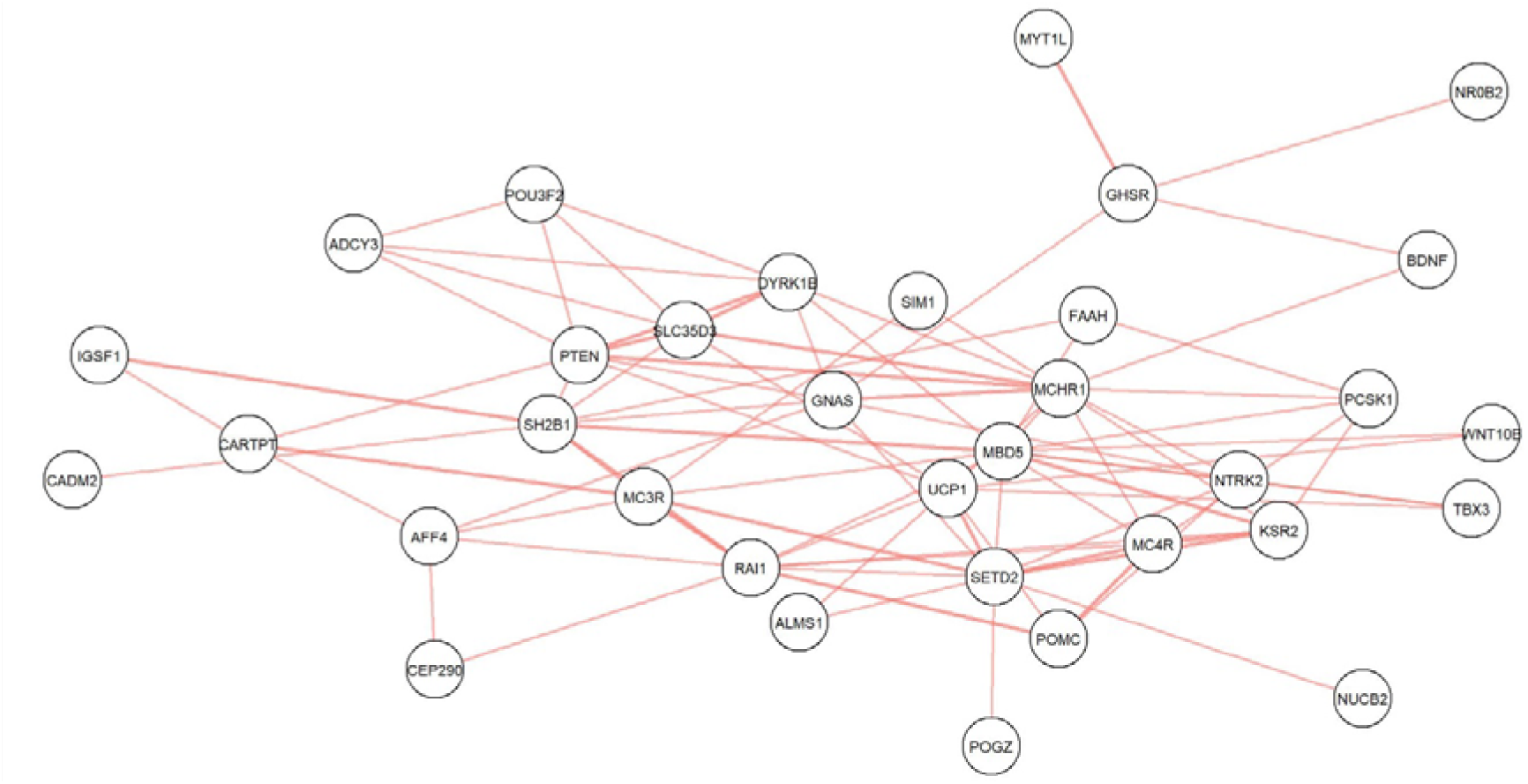
Gene combinations in which PMMO participants carried variants.

## DISCUSSION

In comparison to severe childhood obesity, extreme adult obesity (the mean BMI of PMMO participants is >46) remains almost unexplored genetically and there is no provision for routine testing and genetic counselling, even though therapeutic agents specific to monogenic obesity are available [27]. Here, we aimed to assess the prevalence of variants potentially causing Mendelian forms of obesity in a clinical obesity cohort using a custom-designed rare-variant genotyping array. Since previous analyses in the UKB revealed problems with genotyping rare variants [28], we used an optimised algorithm for analysis [29], visually inspected cluster plots, and included additional samples for quality control. There was good concordance for both samples for which there were duplicates and for samples for which WES data was available.

At least one in twelve PMMO participants have rare variants in genes used by the NHS for a clinical indication of severe childhood obesity [13], increasing to over one in four when including variants in genes on our extended gene list. Since this was intended as a research study (rather than a diagnostic service) and since severe adult obesity is still relatively unexplored in genetic terms, we did not use standard clinical genetics criteria to “prune” mutations for inclusion in our analyses, but 10% would have been classified as variants of uncertain significance (VUS). Our prevalence of possible Mendelian obesity is higher than rates reported by other researchers, possibly because these previous reports considered variants from fewer genes. When including only variants in assigned to genes from an NHS panel typically used for clinical indication of severe early-onset obesity [13], our estimate of 8.6% is closer to previously-reported estimates [4, 5]. In accordance with other studies, 0.88% of PMMO participants carried one or more rare variants in the MC4R gene (excluding the rs2229616 gain-of-function variant) [16].Two PMMO participants carried variants in both the *GHSR* gene and the *MYT1L* gene. Further investigation is needed to determine whether they (or rather their products) interact with one another to influence obesity risk.

We compared our results in the clinical obesity cohort to the UKB whole exome sequence dataset (n = 47,697) and found, to our surprise, that the overall prevalence of qualifying variants was not significantly different, despite the fact that the UKB exhibits “healthy volunteer bias” [12]. Three potential explanations present themselves:

1. Our study design includes investigation of the PMMO participants using a custom rare-variant genotyping array, which has the limitation that other variants that were not reported in the gnomAD dataset (and, therefore, not included in the chip design) may be present in our patients, but undetected – it may be that there is a particular class of variants that were, thus, excluded from the analysis altogether. Exome or whole genome sequencing, and full copy number variant analysis, is required for future analyses.
2. Different variants may have different directions of effect, some increasing risk of obesity and some protecting from it. The majority of the variants in PMMO and UKB participants have not been investigated for function, and it is possible that there is a different distribution of protective genetic factors between the two cohorts. In *MC4R*, the best studied monogenic obesity gene, both loss-of-function (causative of obesity) and gain-of-function variants (pre-disposing to lower BMI) do exist [16] and are not easily distinguished by CADD-PHRED score: for example, rs2229616 has a CADD-PHRED score of 18.5. As seen in our dataset with rs2229616, it may be that the UKB is enriched for the obesity protective variants (it has a well-recognised healthy-volunteer bias [12]) and PMMO participants have more obesity-risk variants. It is difficult to believe that other protective genetic factors do not exist: the prevalence of these may well differ between the two cohorts.
3. The problem of incomplete penetrance: the phenotype may only present if an individual has more than one predisposing risk factor (or fewer protective factors). This could take the form of rare variants in more than one gene (oligogenic inheritance); interaction of rare and common genetic variants, or gene environment interaction. Evidence from previous reports of oligogenic inheritance in other diseases, including autism, cardiovascular diseases and Bardet-Biedel syndrome and our own previous data [7-10], strongly suggests oligogenic inheritance as a possibility. The results presented here appear to support this: PMMO participants with a putatively Mendelian form of obesity were more likely to carry predicted-deleterious mutations in more than one gene than UK Biobank participants.

Our study has several advantages. The PMMO is selected on the basis of an extreme phenotype, i.e. clinical severe obesity. This allows us to explore the prevalence of rare genetic variants that may contribute to clinical obesity with greater statistical power compared to doing this study in a general population cohort and allows us to more accurately estimate the prevalence of rare variants in genes implicated in causing Mendelian forms of obesity. Furthermore, using a custom genotyping array may provide a way for researchers and clinicians to do so even with limited resources.

Despite the cost-effectiveness and time-efficiency of the array, this genotyping methodology has limitations. In particular, we were not able to detect any novel variants because these would not have been present in gnomAD or in HGMD and so were not included in the array design: undoubtedly there will be variants present in the PMMO participants that we have missed. Additionally, the pathogenicity of many of the variants included in the array is not known due to a lack of functional studies, as well as the inclusion of people with obesity in “control” samples. This – along with the fact that new genes, such as PHIP [30], that have been implicated in obesity since the design of the array – limits the accuracy of our estimate of the prevalence of possible Mendelian forms of obesity in our cohort. Lastly, we note that we cannot accurately assess compound heterozygosity, since it is not possible to assign phase, which may mean that we are likely to be underestimating the prevalence of potentially oligogenic forms of obesity in our cohort.

In conclusion, we provide evidence that genetic analysis of adults with severe obesity reveals an underserved population who might benefit from genetic investigation, genetic counselling and targeted therapeutic intervention, and that oligogenic inheritance may play a greater role in severe obesity than previously appreciated. Additionally, our results indicate that – with the recent improvement in calling algorithms – genotyping arrays may be a suitable option for rare variant screening when sequencing is not an option.

## Data Availability

All data produced in the present study are available upon reasonable request to the authors

## Notes

### Competing Interest Statement

The authors have declared no competing interest.

### Funding Statement

Sumaya Almansoori was funded by the Ministry of Higher Education, United Arab Emirates.

### Author Declarations

Ethical approval for the collection and use of data for the PMMO study was given by the NRES Committee London Riverside (REC reference 11\LO\0935) and the research was performed in accordance with the principles of the Declaration of Helsinki. All participants gave informed consent. Also, This research was also approved by Brunel University London Ethics committee application number 26158.

