## Appendix 1 for "Severe obesity may be an oligogenic condition: evidence from 1,714 adults seeking treatment in the UK National Health Service"

There were 4 variants with MAF >5% for which two probes were used to genotype them.

| Variant | Decision | Original probe | Alternative probe |
| --- | --- | --- | --- |
| 2:169140540:C:T | Alternative probe. No carriers. | 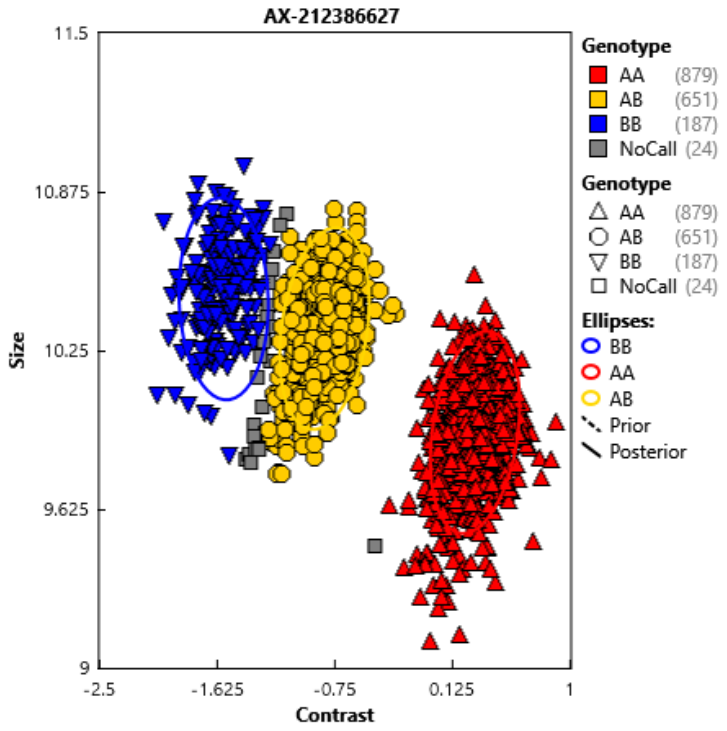 | 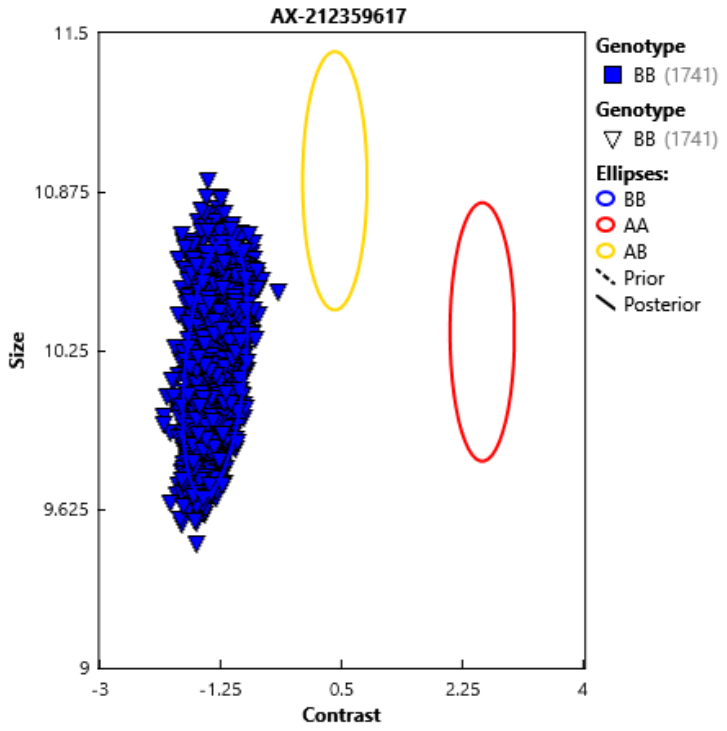 |
| 6:100420902:G:A | Alternative probe. No carriers. | 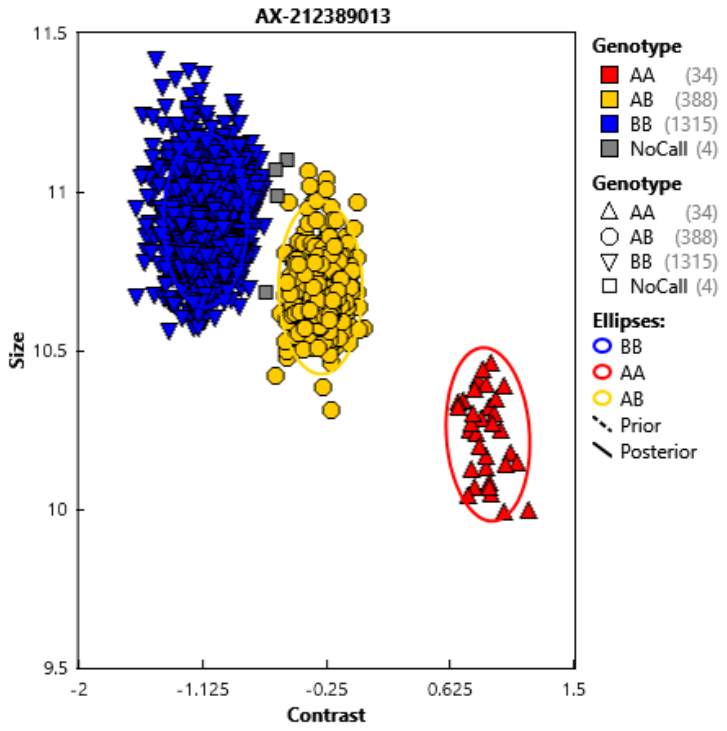 | 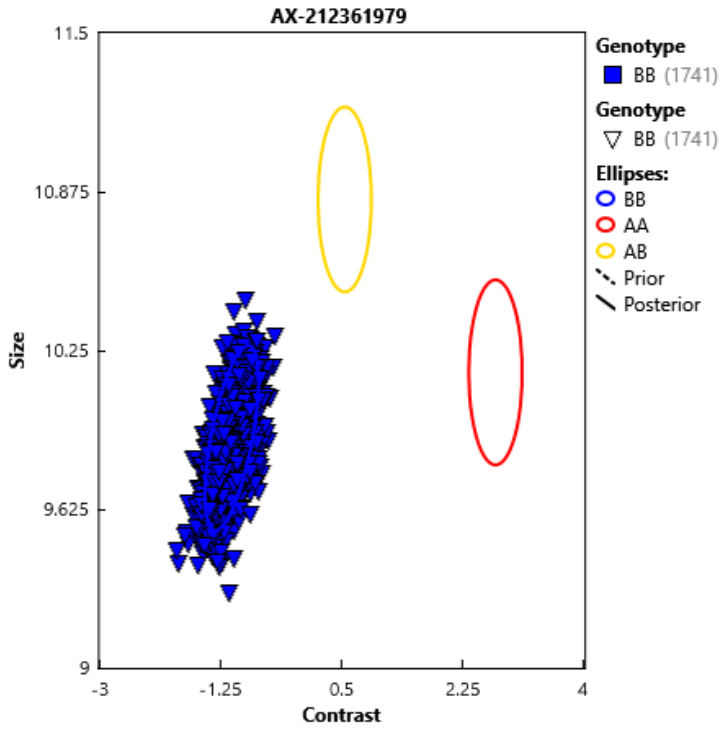 |
| 12:117761480:TC:T | Excluded due to presence of common variant (rs617641 12:117761481:C:G) immediately downstream | 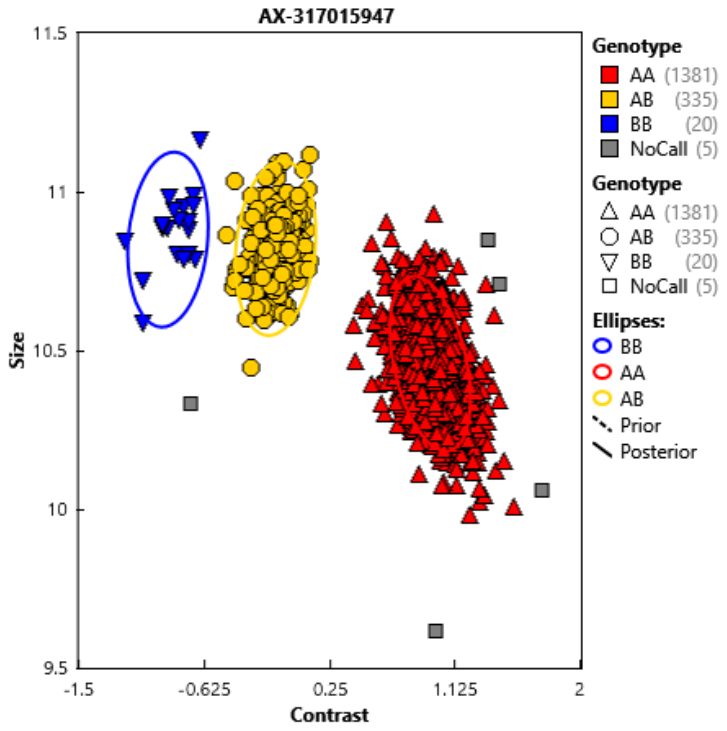 | 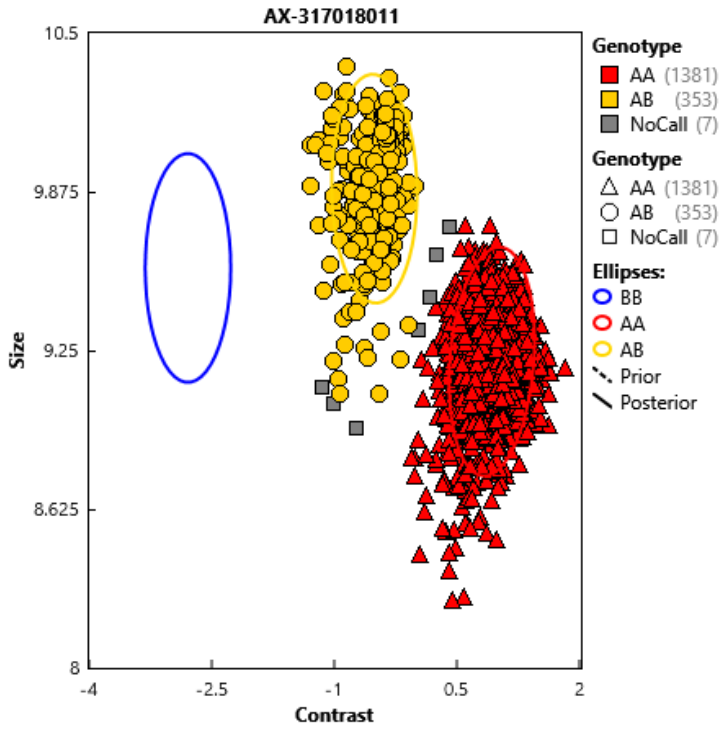 |
| X:131282718:C:G | Be conservative and assume no carriers because plots are inconsistent | 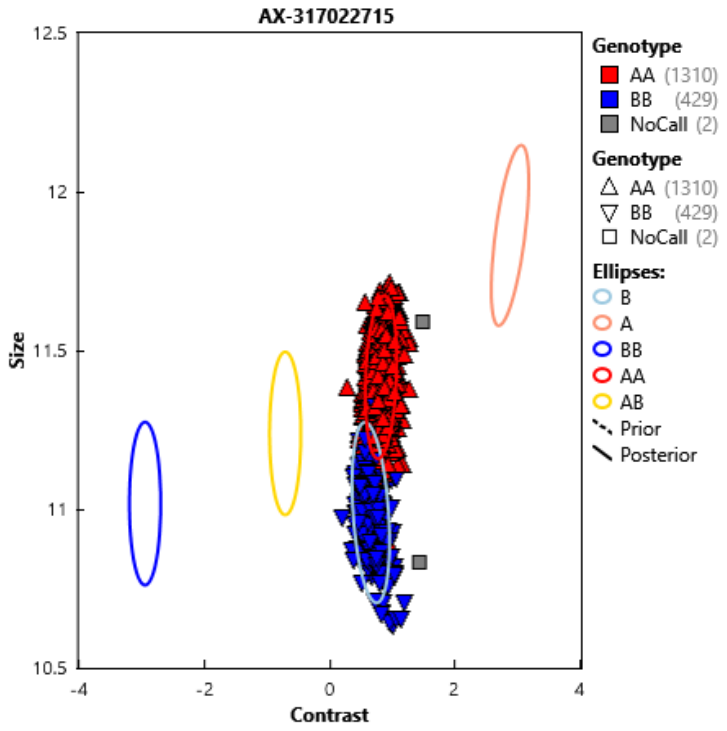 | 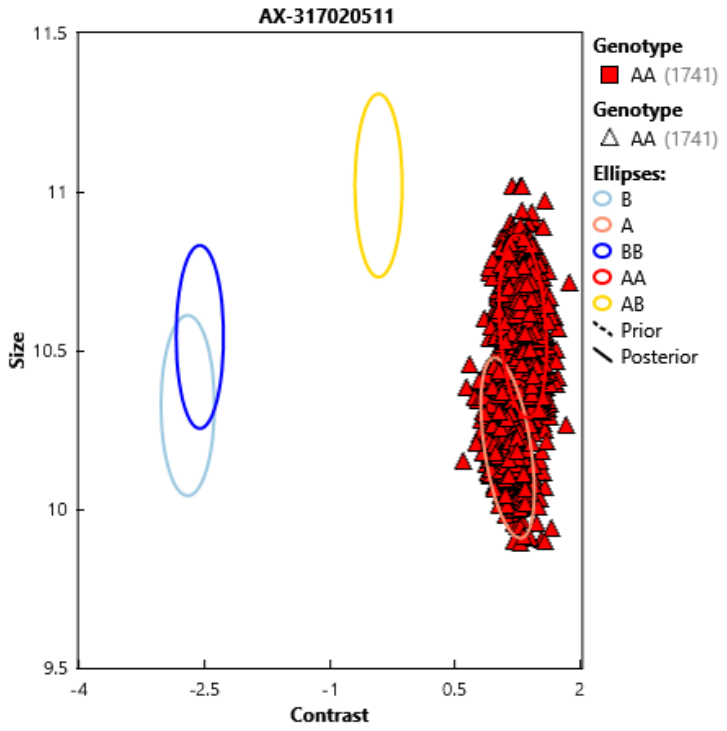 |
| 20: 58909185:TC:TG | Exclude because cluster plots do not look optimal and there is a common variant (rs8620 20:58909186:C:T) at the same locus that the probe is targeting | 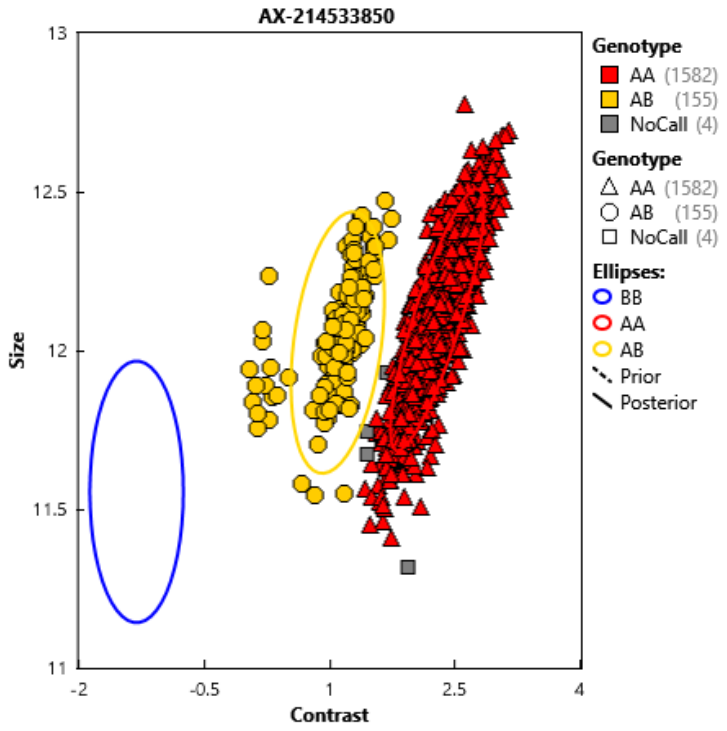 | 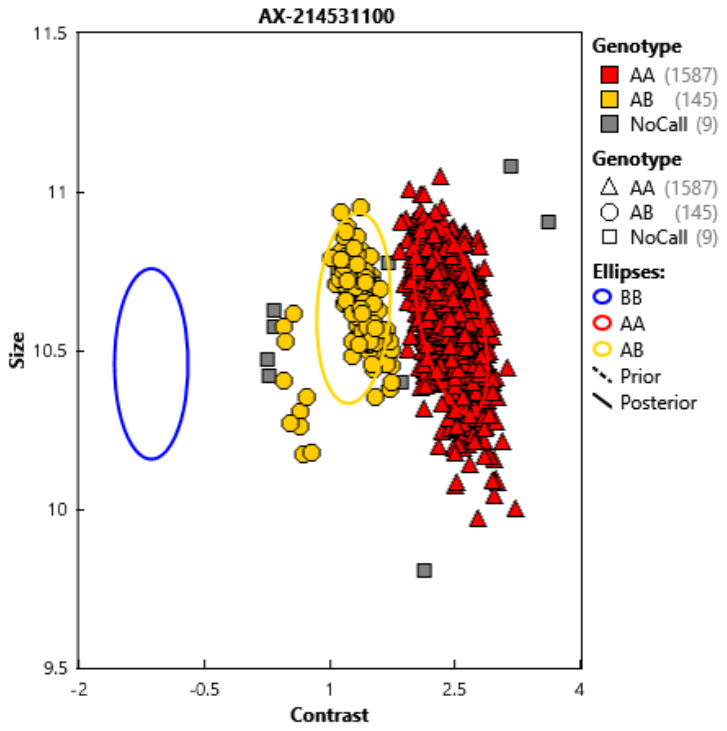 |

Variants with a MAF >5% for which there was only probe used

| Variant | Decision | Probe |
| --- | --- | --- |
| 4:122743048:CG:TA | Excluded due to presence of common variant (rs309370 4:122743049:G:A) immediately downstream | 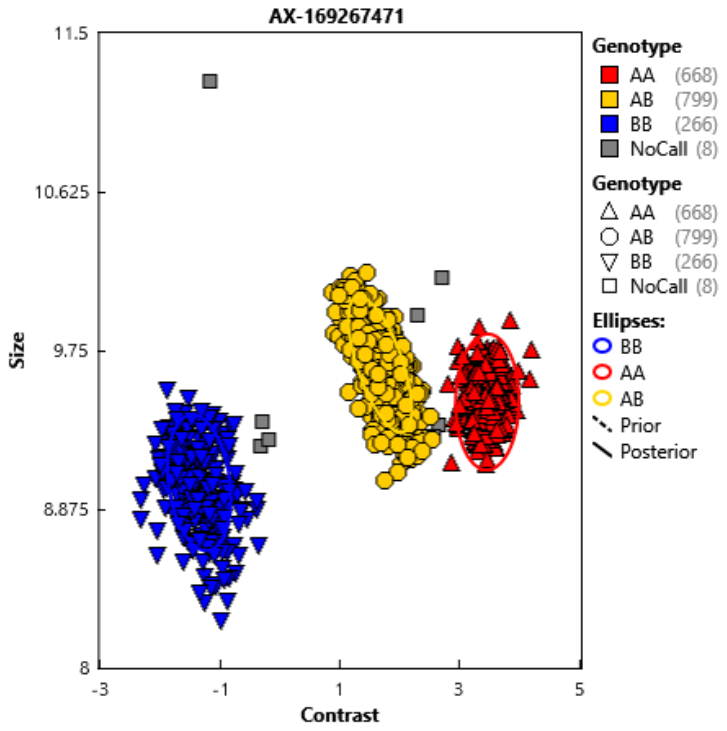 |
| 2:73453381:G:T | Excluded because clusters are not separated well enough and the locus is also multiallelic (gnomAD v3.1.2 AF of 2:73453381:G:C is 0.387 whereas gnomAD AF of 2:73453381:G:T is 6.59e-06) | 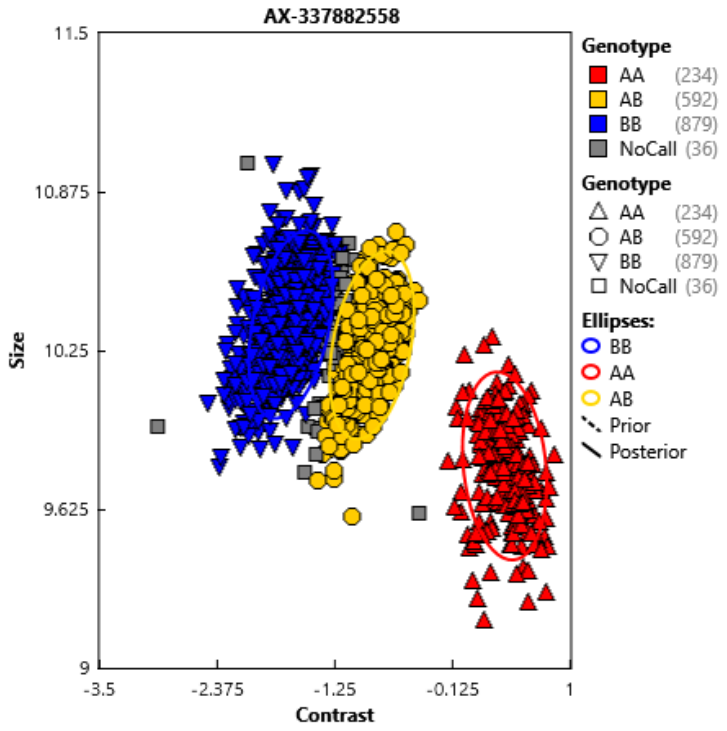 |
| 6:100420903:G:A | Excluded because the locus is multiallelic (gnomAD v3.1.2 AF of 6:100420903:G:T is 0.129 whereas gnomAD AF of 6:100420903:G:A is 1.51e-04) | 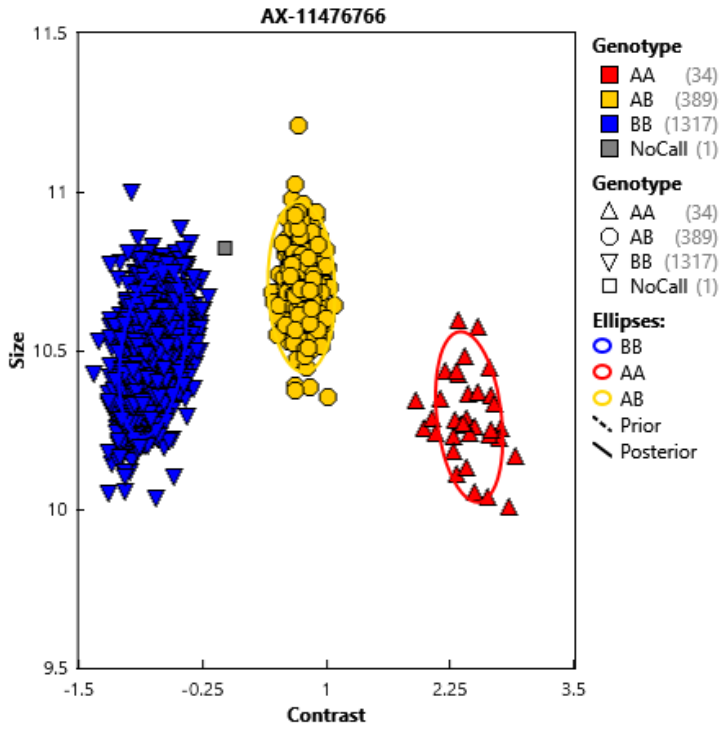 |
| 2:73489683:G:C | Excluded because clusters are not well separated and locus is also multiallelic (gnomAD v3.1.2 AF of 2:73489683:G:A is 0.124 whereas gnomAD AF of 2:73489683:G:C is 1.316e-04) | 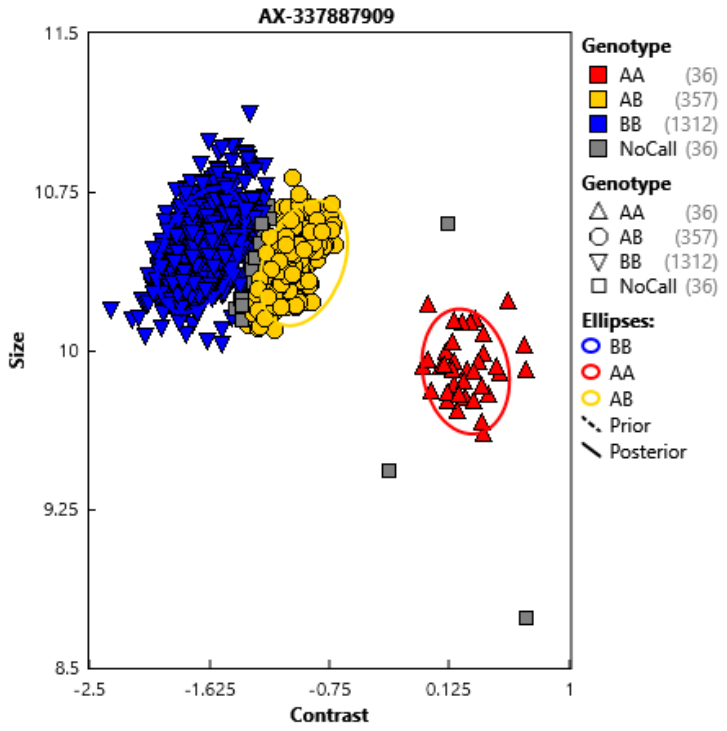 |
| 12:117761473:G:C | Clusters are well separated, no common alternative allele at this locus and no common variant immediately upstream/downstream. However, this variant was excluded due to an outlying minor allele count | 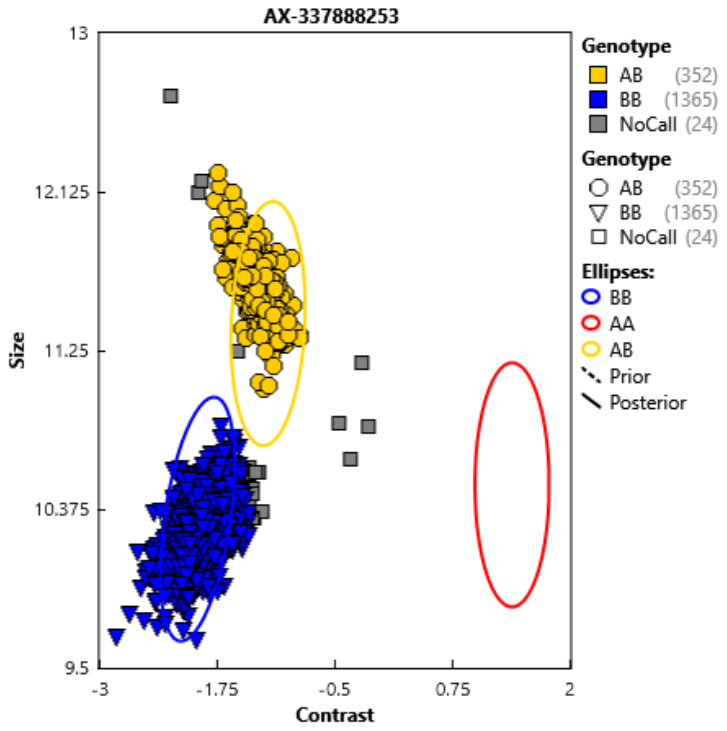 |
| 15:72727469:G:C | Keep because clusters are well separated and no evidence in gnomAD of common alternative alleles nor of common variants immediately upstream/downstream | 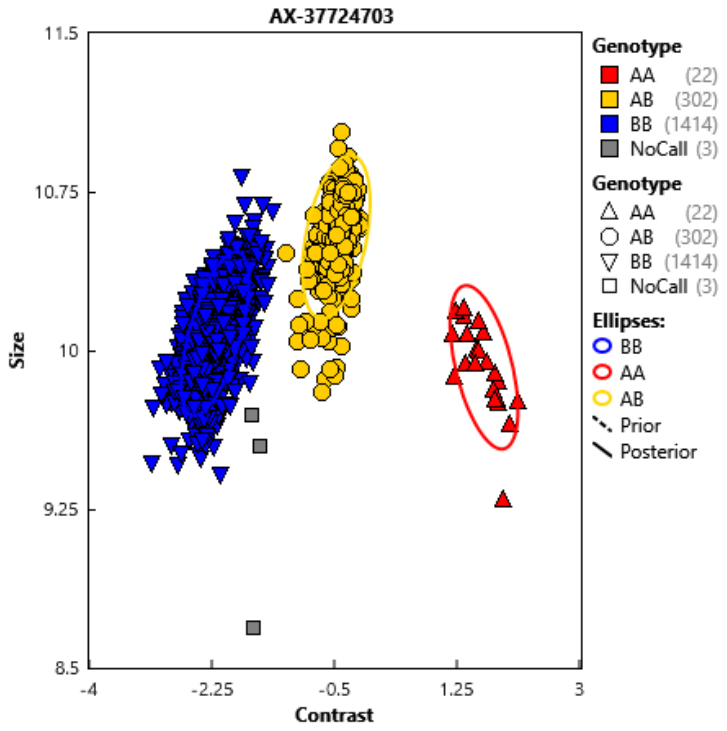 |
| 22:40679491:C:A | Exclude because clusters not well separated | 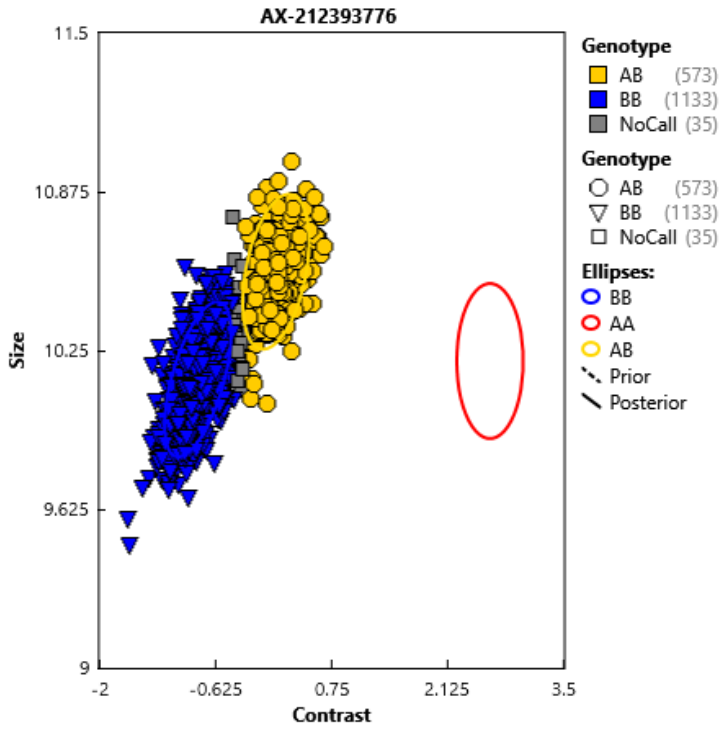 |

Variants that are PolyHighRes for which two probes were used to genotype them

| Variant | Decision | Original | Alternative |
| --- | --- | --- | --- |
| 20:58909185:TC:T | There appears to be no meaningful discrepancy between the two probes. However, this variant was excluded due to an outlying minor allele count | 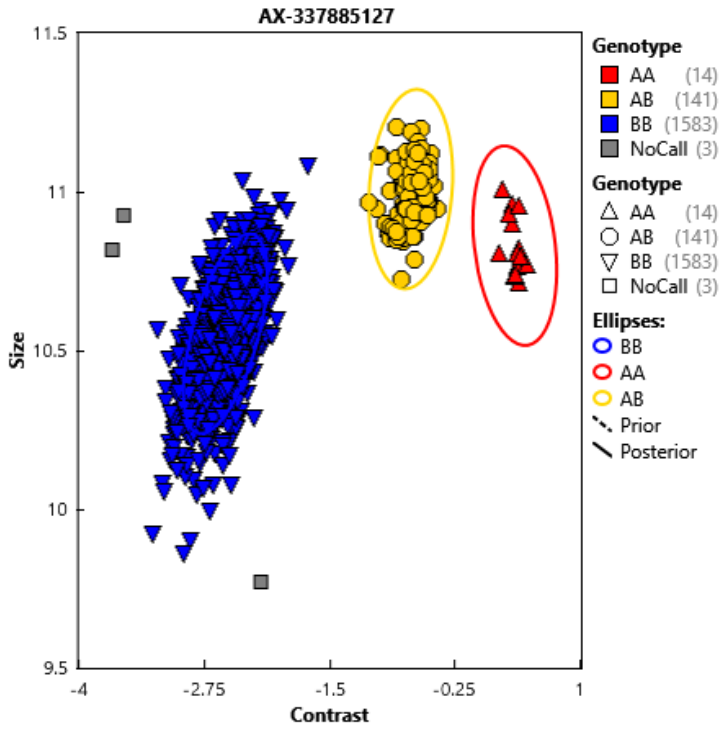 | 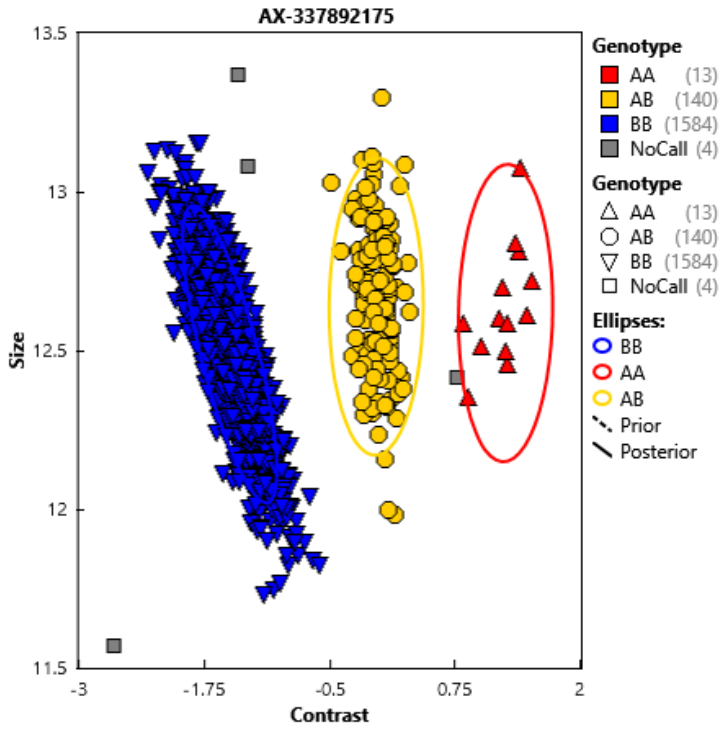 |
| 2:63439843:G:A | Alternative probe. No carriers. | 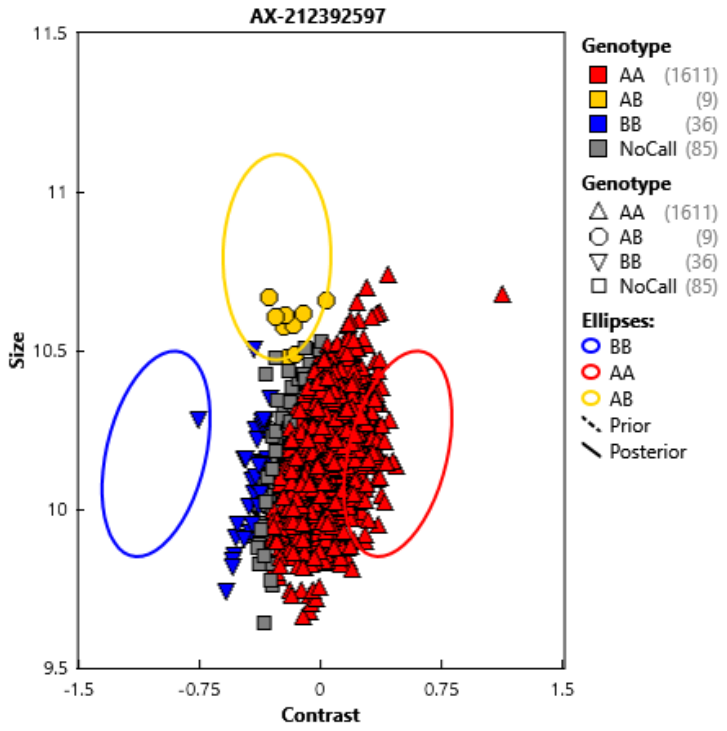 | 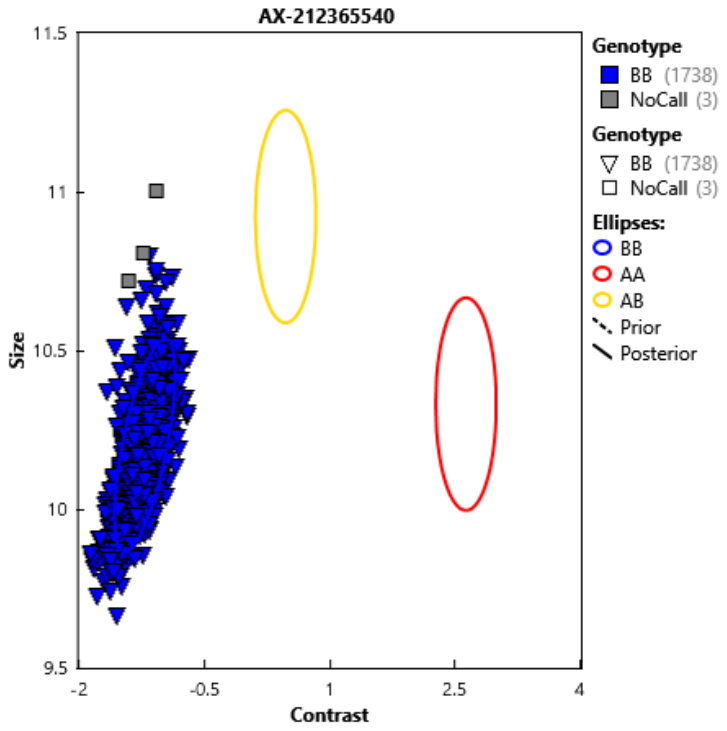 |
| 2:1922844:C:T | Alternative probe. No carriers. | 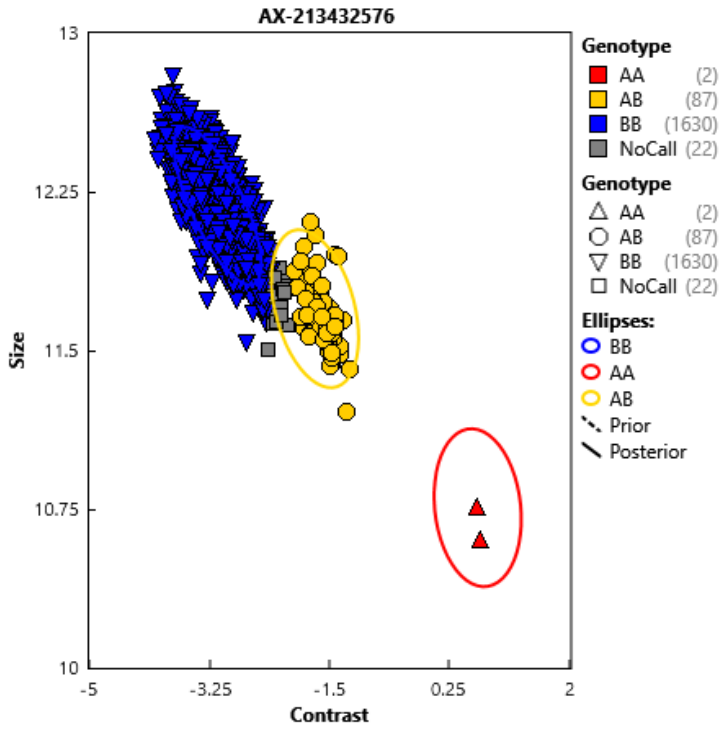 | 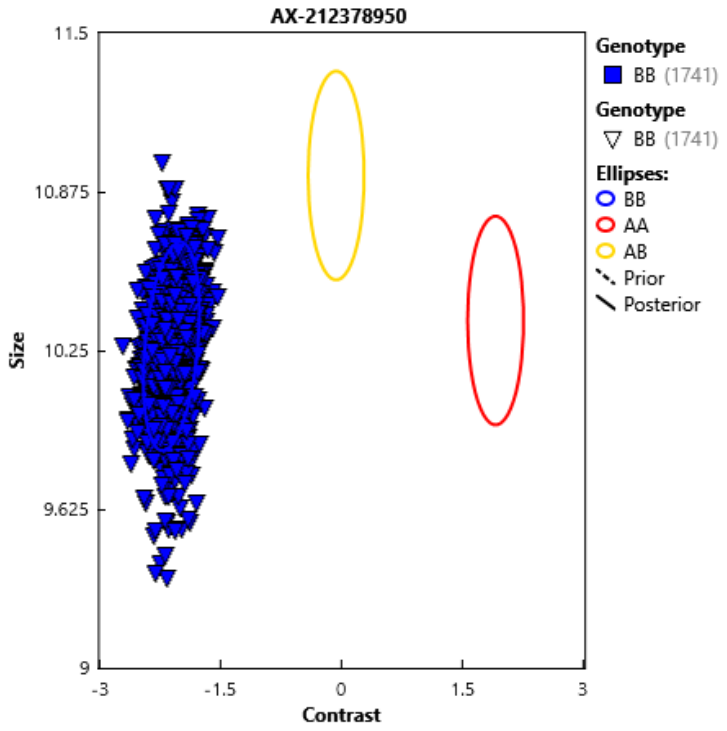 |
| 1:65616108:C:T | Original probe | 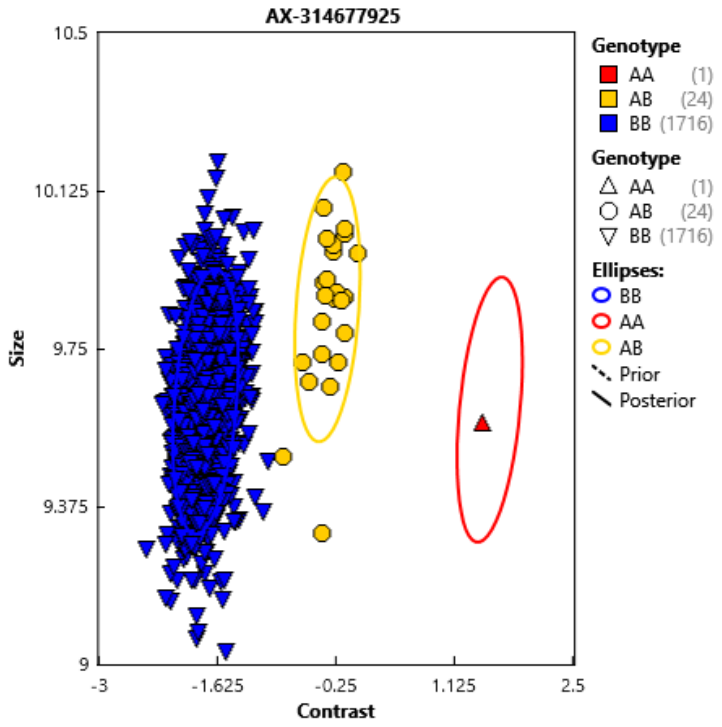 | 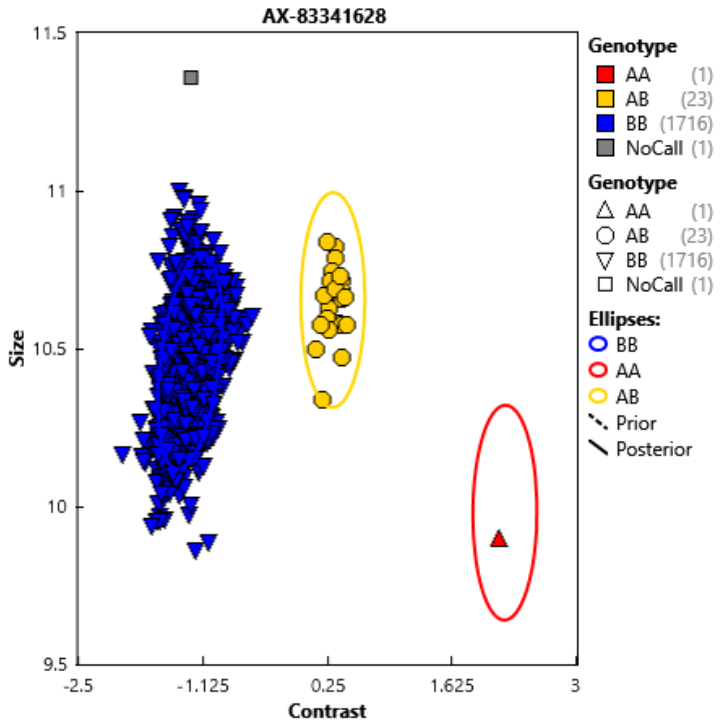 |
| 12:88086456:C:G | Original probe | 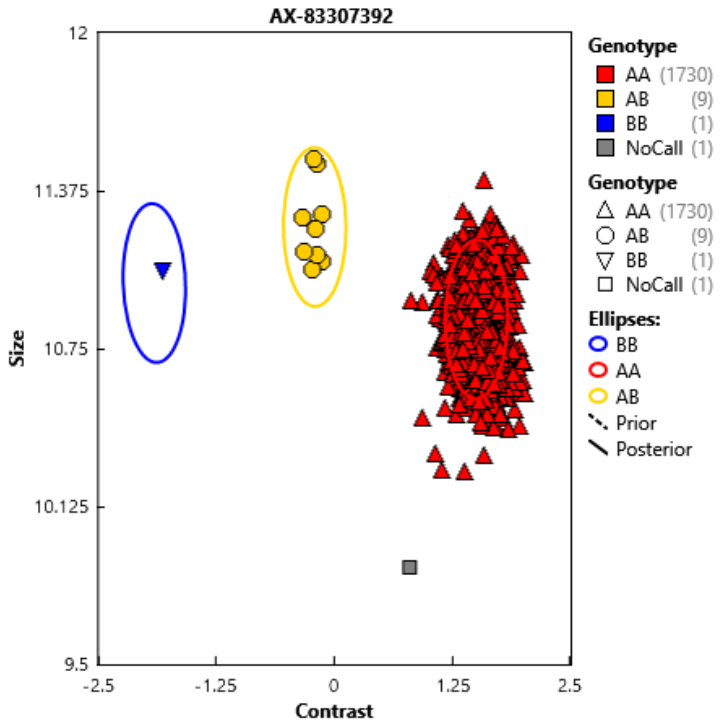 | 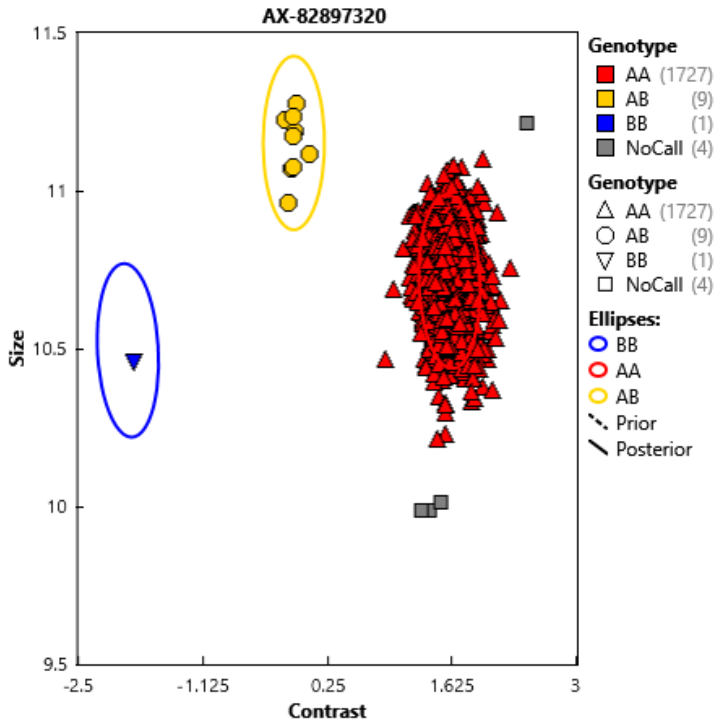 |
| 15:72731341:G:A | Original probe | 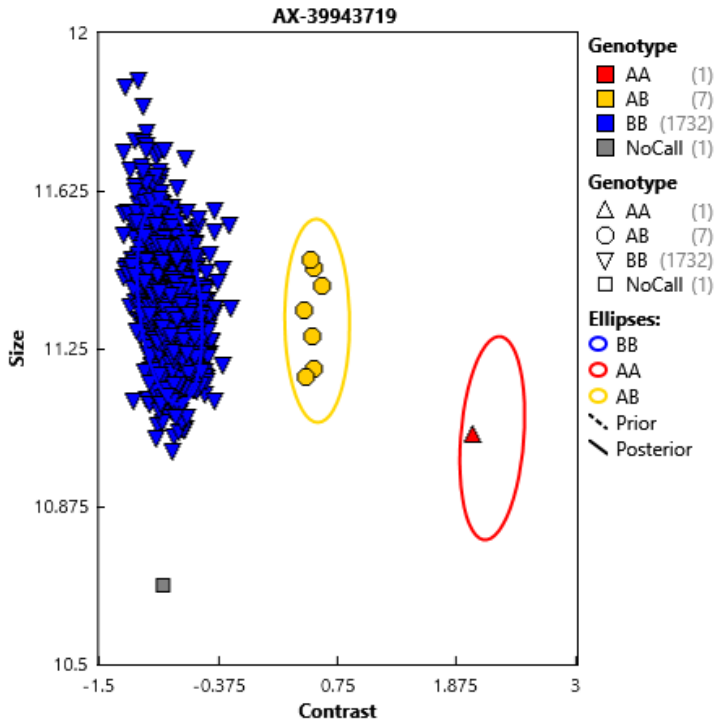 | 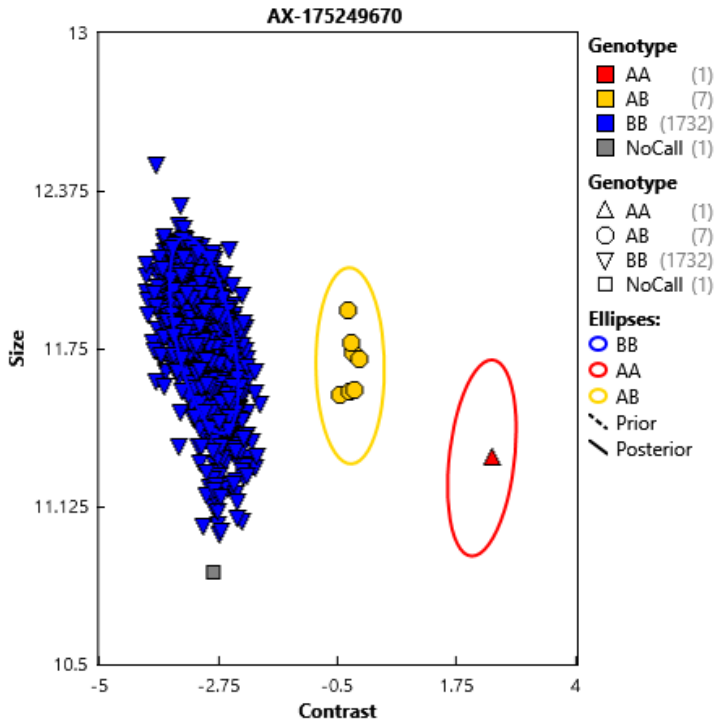 |
| X:131274090:G:A | Original probe | 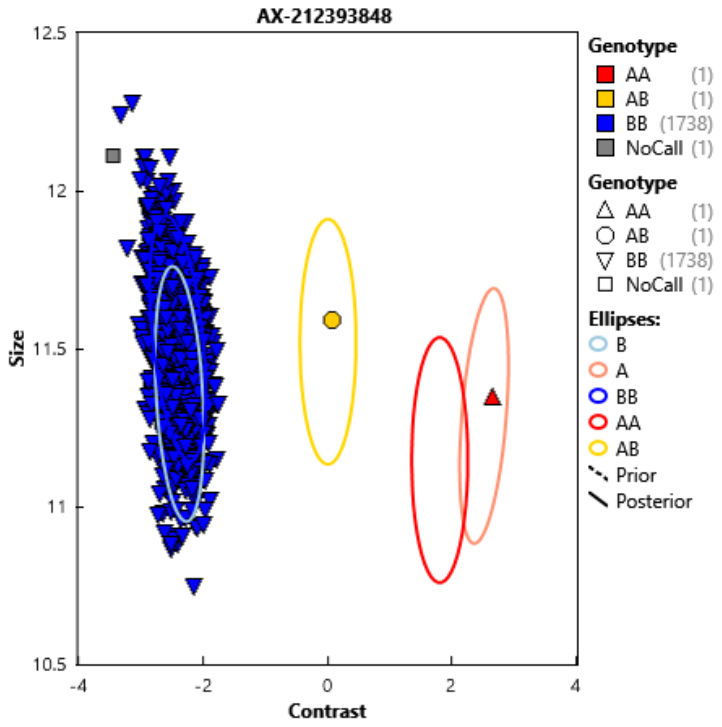 | 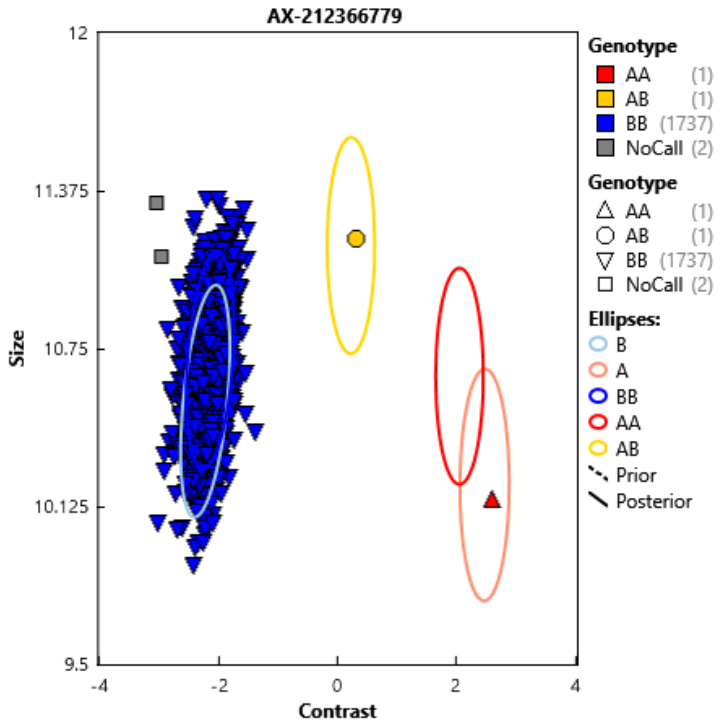 |
| 1:151428125:A:T | Alternative probe. No carriers. | 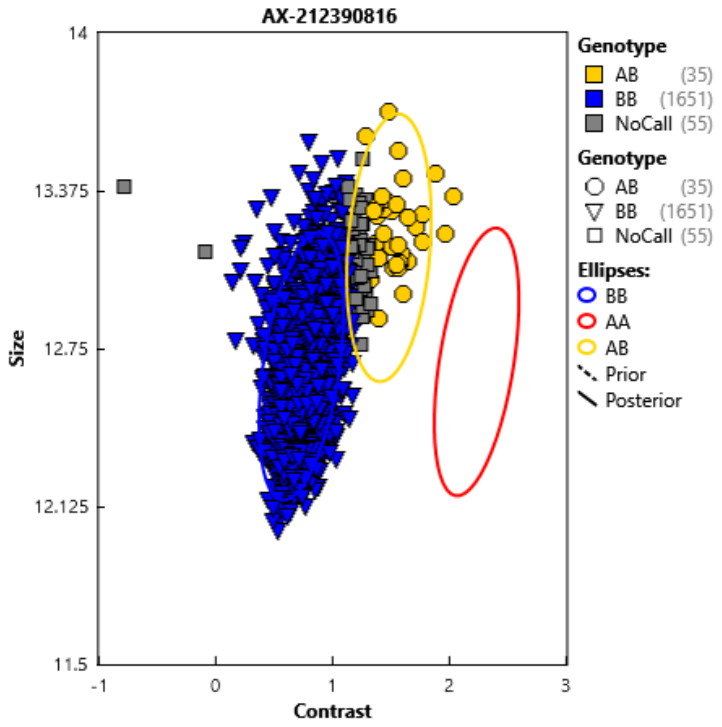 | 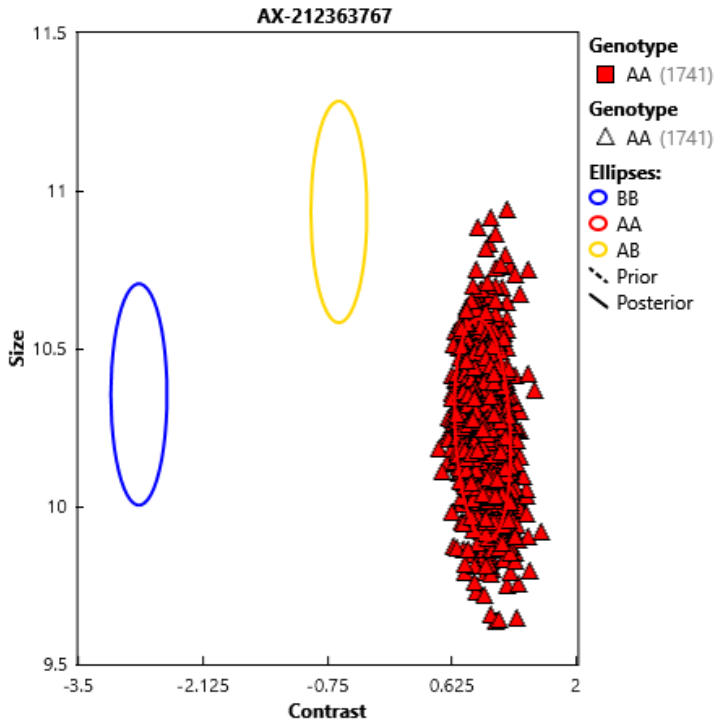 |
| X:131285137:G:C | Original probe | 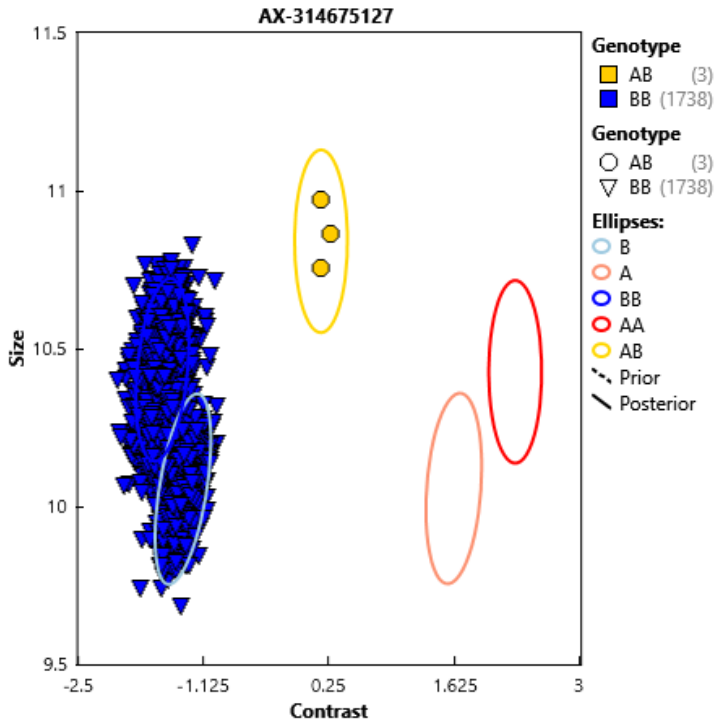 | 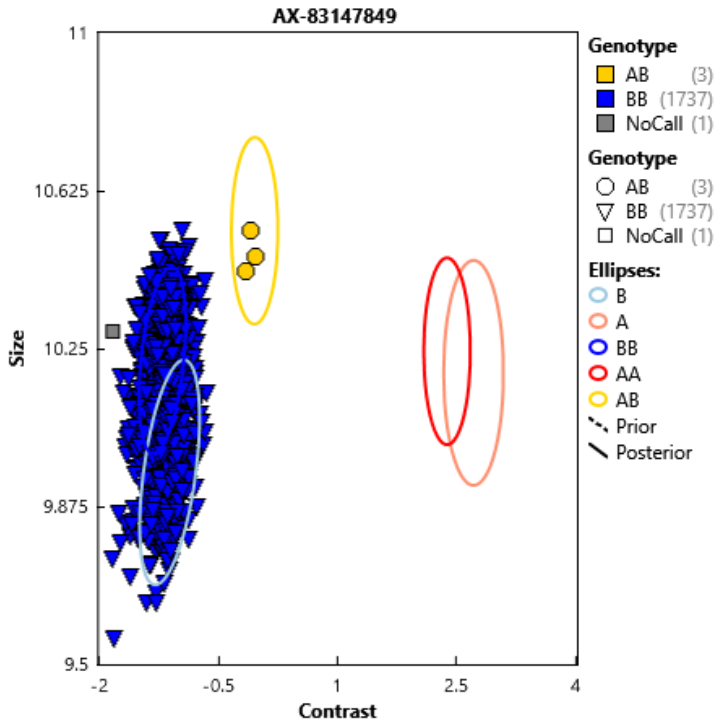 |
| X:131278596:G:T | Original probe | 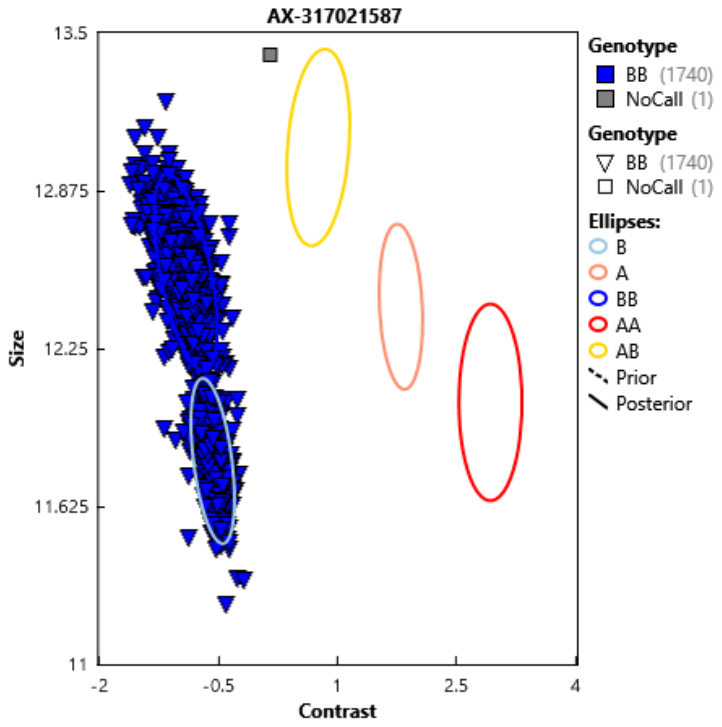 | 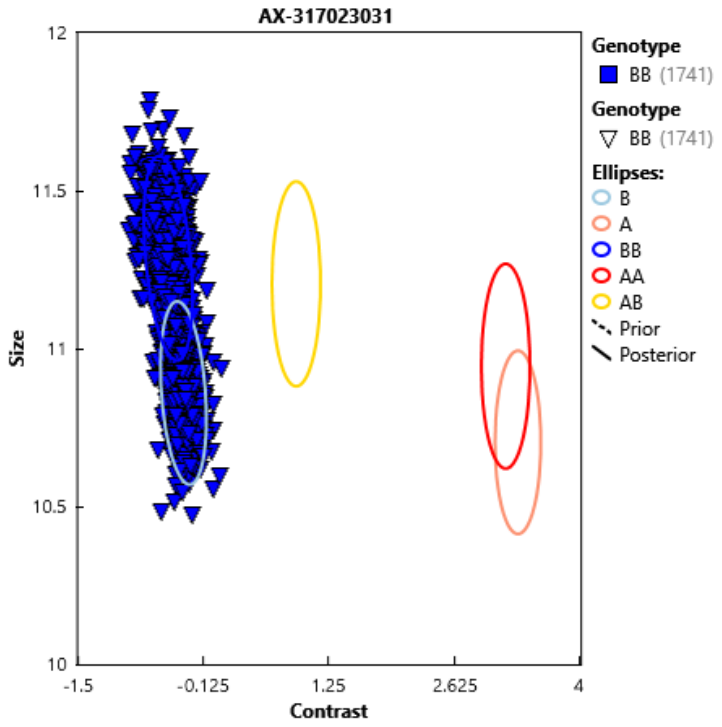 |
| X:131278575:C:T | Original probe | 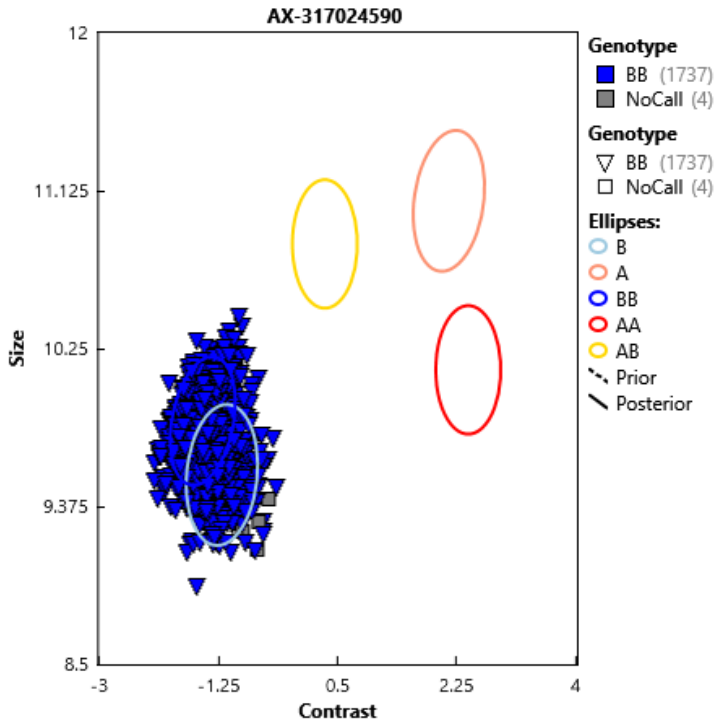 | 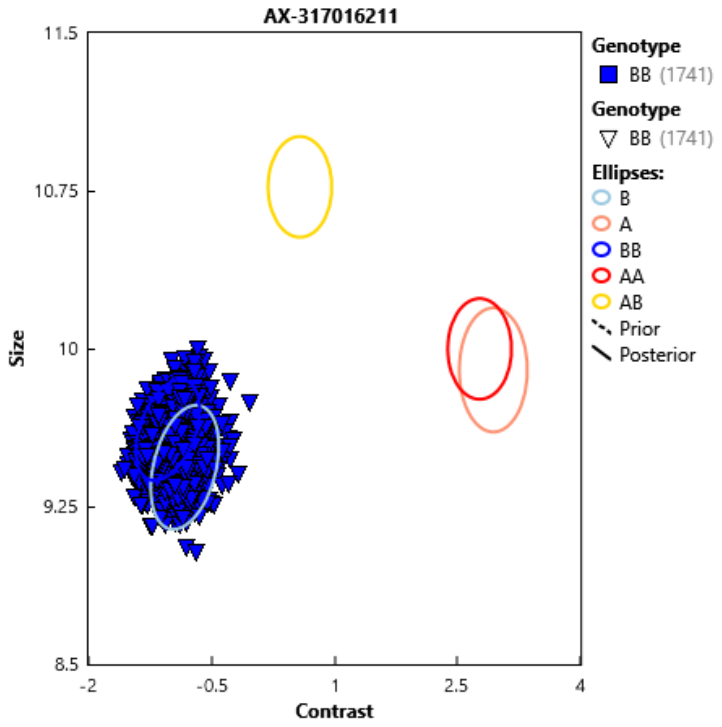 |

Variants that are PolyHighRes with one probe

| Variant | Decision | Probe |
| --- | --- | --- |
| 1:65534922:T:C | Exclude because clusters are not well separated | 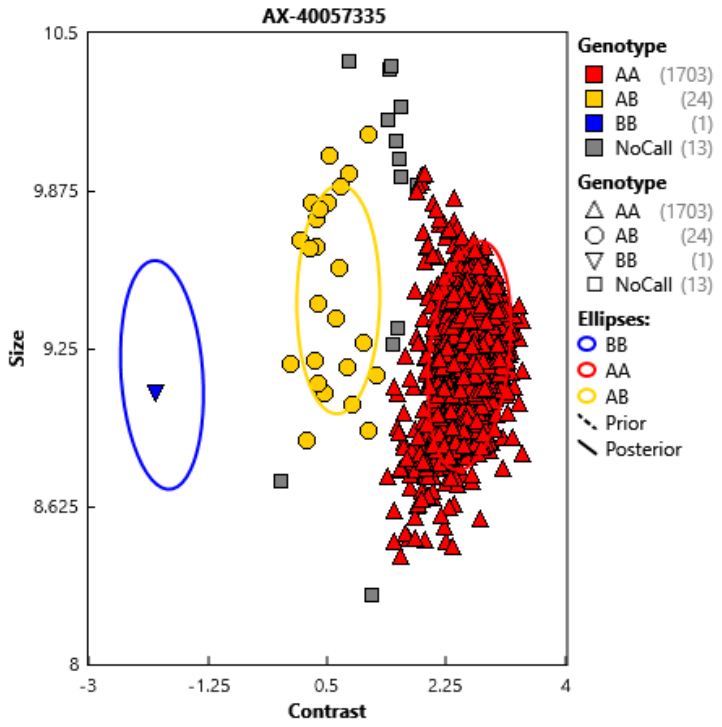 |
| 2:1923182:T:A | Exclude because clusters are not well separated | 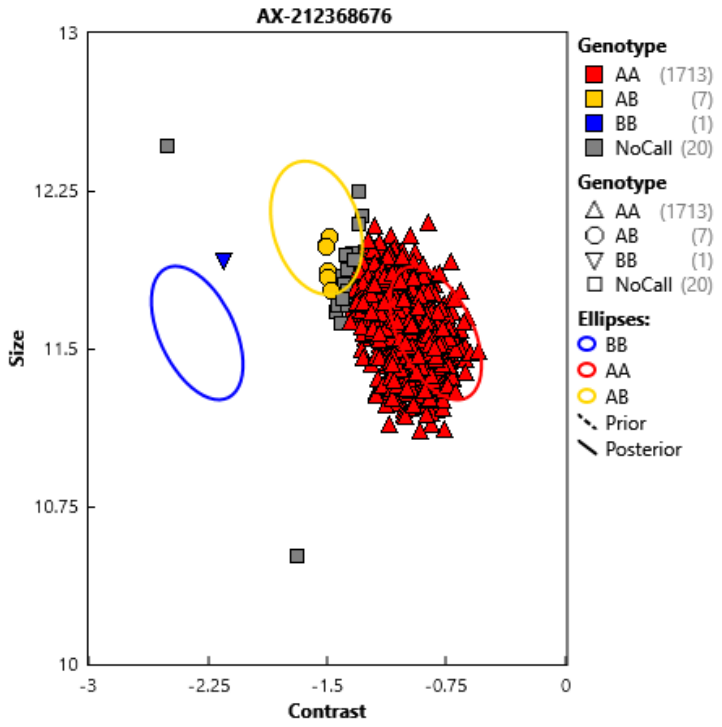 |
| 16:28873537:C:T | Keep | 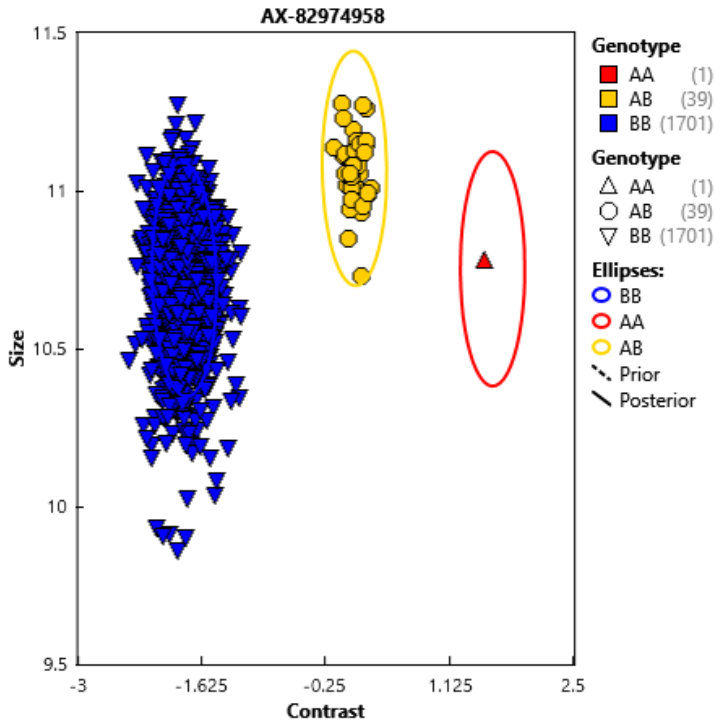 |
| X:131278706:T:G | Keep | 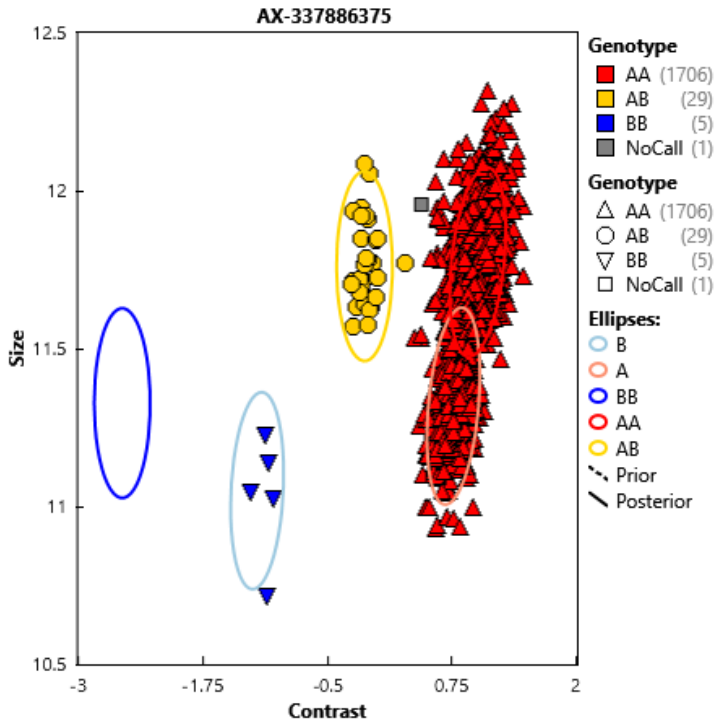 |
| 20:10412791:C:A | Keep | 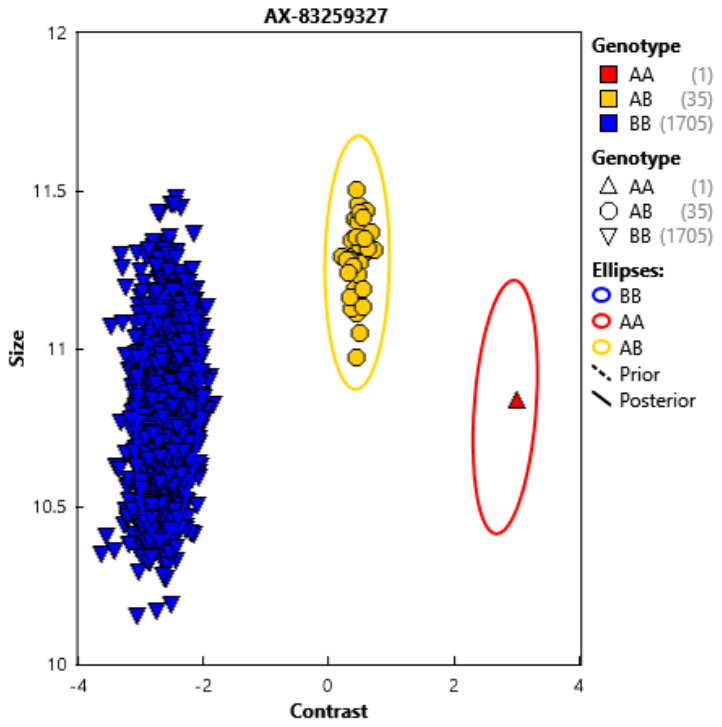 |
| 18:60371598:ATC:AGC | Keep | 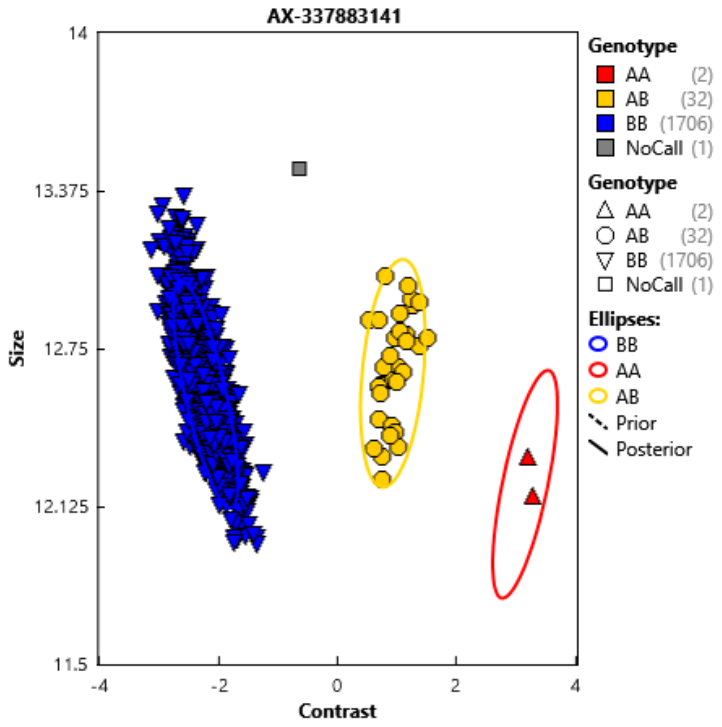 |
| 2:73572505:C:G | Keep | 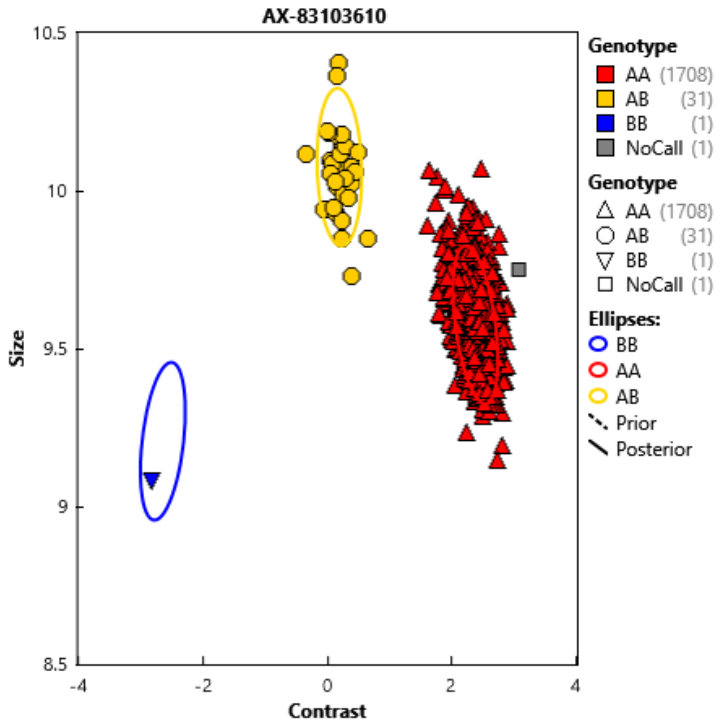 |
| X:131275723:A:G | Keep | 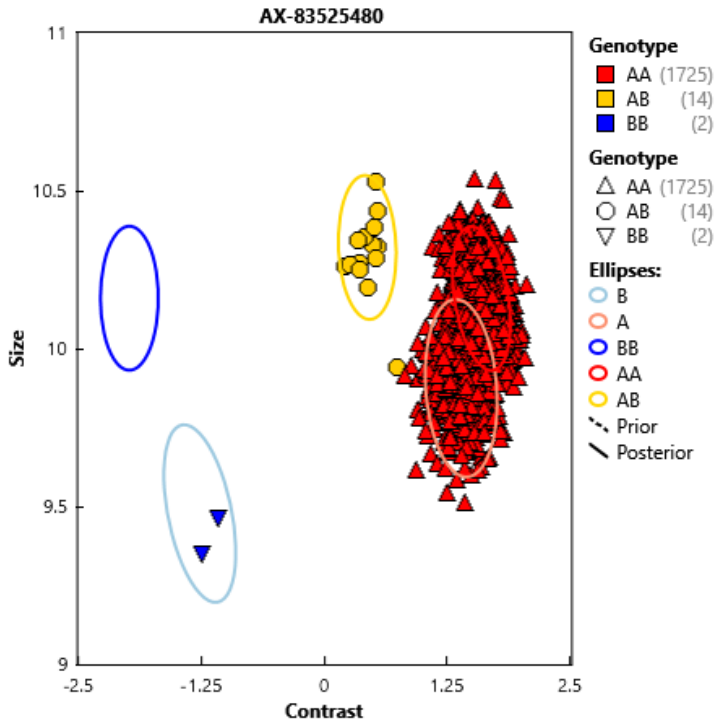 |
